# Replacing cars with green spaces: an assessment of the mortality benefits in Paris

**DOI:** 10.1101/2025.07.21.25331925

**Authors:** Léo Moutet, Lucie Adélaïde, Charles Claron, Kamel Bahri, Mohamed Ali Ben Halima, Johanna Lepeule, Mathilde Pascal, Laura Temime, Kévin Jean

**Affiliations:** MESuRS Laboratory, Conservatoire national des Arts et Métiers (Cnam), Paris, France; Paris Research in Health Environment and Climate (PARSEC), Ecole Normale Supérieure, Inserm,Paris, France; Université Grenoble Alpes, INSERM U1209, CNRS UMR 5309, Institut pour l’Avancée des Biosciences (IAB), Team of Environmental Epidemiology Applied to Development and Respiratory Health, 38000 GRENOBLE, France; Santé Publique France, Department of Environmental and Occupational Health, French National Public Health Agency, Saint Maurice 94415, France; CIRED, École des Ponts, AgroParisTech, EHESS, CIRAD, CNRS, Université Paris-Saclay, Nogent-sur-Marne, France; Employment and Labour Research Centre (CEET), Conservatoire national des Arts et Métiers (Cnam), Paris, France; Eco-Evolutionary Mathematics team, IBENS, École Normale Supérieure, CNRS, INSERM, Université Paris Sciences & Lettres, Paris, France

## Abstract

Increasing urban vegetation coverage is associated with improved human health and well-being, reduced environmental impact of cities and enhanced urban resilience to climate change. To support evidence-based urban planning, this study quantifies the mortality benefits, equity implications and cost-benefit ratio of several scenarios of green space development in Paris by 2040, including the replacement of car-dedicated surfaces with green spaces and a best-case scenario. This quantitative health impact assessment is based on estimated changes in the Normalized Difference Vegetation Index (NDVI), obtained through the estimation of the dynamic effects over time using a Difference-in-Differences approach based on previous public greening interventions, and on an exposure-response relationship linking NDVI and all-cause mortality. It was conducted at the sub-municipal level (IRIS) and incorporates a social deprivation index to assess health equity implications. Vegetation costs are drawn from a previous French study estimating urban soil restoration prices. Replacing surplus on-street parking and 20% of street space with vegetation could reduce all-cause mortality by around 0.8%, while reaching 15% of vegetation coverage in each IRIS could prevent around 3% of deaths yearly in Paris as early as 2040. For all scenarios, these benefits were approximatively equally distributed across deprivation levels. Predicted monetised health benefits outweigh intervention costs by 2035, with further impacts representing net gain. In conclusion, greening interventions targeting car-dedicated space in Paris would equitably improve health while supporting more sustainable and resilient cities.

## Introduction

Urban green spaces (UGS) enhance health and well-being, for instance by improving mental health, pregnancy outcomes, reducing cardiovascular morbidity, prevalence of type 2 diabetes or all-cause mortality (Ahmer et al., 2024; Shrestha et al., 2024; Wang et al., 2025). Exposure to green spaces, in addition to reducing exposure to environmental stressors (air pollution, traffic noise or urban heat island effect), is associated with capacity developments (increased physical activity and social cohesion), mental recovery (cognitive restoration, reduced stress) and improved immune system functions (Markevych et al., 2017; WHO Regional Office for Europe, 2016). While green spaces can also introduce certain risks such as allergen exposure or vector-borne diseases, the overall epidemiological evidence supports their role in enhancing population health (Rojas-Rueda et al., 2019; WHO Regional Office for Europe, 2016). UGS often confers a greater protective effect to lower socio-economic groups; however, the potential increases in land value and influx of higher-income residents may trigger green gentrification, increasing health disparities in access to and benefits from UGS. (Anguelovski et al., 2022; WHO Regional Office for Europe, 2016).

UGS, in addition to promoting health, also appear as an effective climate mitigation and adaptation strategy in urban areas, providing multifunctional ecosystem benefits despite variability, disturbance and management uncertainty (IPCC (Intergovernmental Panel on Climate Change), 2023; McPhearson et al., 2015). Our societies, especially in dense urban areas, must implement targeted mitigation and adaptation actions to promote sustainable development (IPCC (Intergovernmental Panel on Climate Change), 2023) and provide unconditional short-term health co-benefits to their populations (Moutet et al., 2025). Climate change will increasingly affect our cities, and as about 85% of the European population is projected to live in urban areas in 2050, there is a need to improve cities’ resilience (IPCC (Intergovernmental Panel on Climate Change), 2023; United Nations Department of Economic and Social Affairs, 2019). While flooding and urban heat risks increase, ecosystem-based approaches are recognized as effective adaptation measures for dense cities and contribute to biodiversity conservation (IPCC (Intergovernmental Panel on Climate Change), 2023). In European cities, nature-based solutions are also known to reduce carbon emissions by up to 25% and lower mean city temperature by 0.4°C (up to 1.3°C) during summer, preventing 1.8% of heat-related deaths (Pan et al., 2023).

In this context, the municipality of Paris is implementing a climate action plan aimed at carbon neutrality, with operational adaptation means prioritizing sustainability and nature-based solutions while protecting the most vulnerable populations (Mairie de Paris, 2023). As a member of the C40 Cities network, Paris municipality signed the C40 Urban Nature Accelerator, pledging to reach 30-40% of its surface with green space or permeable surface and ensuring access to UGS for 70% of its population by 2030 (C40 Cities Climate Leadership Group, n.d.). An earlier study showed that reaching the WHO targets on accessible green space would yield important health benefits in several European cities, including Paris (Barboza et al., 2021). However, to our knowledge, no existing framework quantifies changes in NDVI following spatially explicit urban green space implementation, thus allowing the estimation of associated health impacts and a cost-benefit analysis.

Finding available space in dense cities can be a barrier to greening strategies, while in those areas, exceeding space is allocated to cars (Creutzig et al., 2020). Parking and road space are also linked with public health issues such as car dependency (and its associated impact on air pollution, climate and biodiversity), impervious surfaces (responsible for urban heat, flooding and water pollution) and reduced housing affordability, all associated with downstream effects on health and social inequality (Garber et al., 2024; Miner et al., 2024). Transformation of impervious surface to green space implies multiple steps with various costs depending on baseline conditions and restoration strategies (Salin et al., 2025). Assessing the mortality benefits of specific local policies could favour healthy urban planning. Therefore, this study aimed to estimate the mortality benefits of plausible UGS developments, notably by replacing car-dedicated space in Paris, while specifying equity implications and a cost-benefit analysis.

## Methods

We conducted a quantitative health impact assessment of in-house scenarios projecting various greening intervention for 2040. Our study focused on the municipality of Paris, composed of 992 IRIS (*Ilots Regroupés pour l’Information Statistique*), a sub-municipal unit that respects geographic and demographic criteria. We relied on a relative risk (RR) linking all-cause mortality with the Normalized Difference Vegetation Index (NDVI) to estimate mortality benefits from our greening scenarios.

### Exposure to urban green space

We used the Normalized Difference Vegetation Index (NDVI) as a proxy for exposure to UGS given its association with all-cause mortality in a meta-analysis of longitudinal studies. This indicator quantifies vegetation growth by calculating the difference between near-infrared and visible reflectance, divided by their sum, using remote sensing imagery. The NDVI has a range from −1 (water) to +1 (dense vegetation), with values approaching 0 corresponding to a lack of vegetation. We used the NDVI calculated from Landsat surface reflectance with a 30m spatial resolution based on Robinson et al. 2017 (Robinson et al., 2017). Mean summer (June to august) NDVI from 2000 to 2018 was retrieved from previous studies (Adélaïde et al., 2024; Hough et al., 2023) at the IRIS level to be consistent with the geographical population density. Within each IRIS, NDVI was considered homogeneous. IRIS units are continuous, homogeneous, and stable geographic areas defined by INSEE to enable the dissemination of local statistics, usually composed of 1,800 to 5,000 inhabitants. According to our data, IRIS units in Paris have a mean surface area of 11 hectares (range: 1-538) and a mean population of 2 277 residents (range: 0-7572).

### Model calibration for NDVI evolution following greening interventions

To evaluate the effect of spatially explicit greening interventions (as defined in our first and second scenarios) on the NDVI at the IRIS level, we investigated public green space implementations from 2001 to 2017 in Paris (Supplementary figure S1), and measured, for each of them, the corresponding change in NDVI. We then computed a calibration factor, defined as the ratio of the absolute NDVI change relative to the proportion of surface area (at the IRIS level) that was converted into a green space. This approach allows to link the number of square meters greened to the NDVI evolution, based on past local dynamics.

To compute the calibration factor from the observed data, we employed the “Difference-in-Differences Estimators of Intertemporal Treatment Effects” approach(De Chaisemartin and D’Haultfœuille, 2024), which allows for the estimation of dynamic causal effects over time, in the presence of staggered and continuous treatment intensities (surfaces greened). This event-study framework was particularly suited for our objective, as it enabled us to capture the temporal evolution of NDVI following greening interventions across multiple spatial units (IRIS) and time periods. Unlike traditional binary-treatment Difference-in-Differences models, this method accommodates a continuous treatment variable—in our case, the proportion of surface area greened within each IRIS—thus reflecting more realistic, gradual intervention intensities. It also allows for heterogeneous treatment timing, accounting for the fact that greening projects were not implemented simultaneously across the city. Using the “did_multiplegt_dyn” package in R, we computed the average cumulative effect (i.e., NDVI change over time) of the treatment (greening interventions) relative to the treatment intensity (i.e., the proportion of IRIS greened), with a 95% confidence interval. The key assumptions underlying the model—namely, the absence of anticipation effects and the parallel trends assumption—are tested by the package and were confirmed to hold true (Supplementary Figure S2), thereby validating the robustness of our causal inference. The effect was computed over 14 year-periods to account for lagged treatment effects related to vegetation growth. More details on the calibration are available in Supplementary text 1. Green space implementation data in Paris (creation date, size and location) are publicly available (Direction des Espaces Verts et de l’Environnement - Ville de Paris, 2025).

### Scenarios

We constructed three plausible scenarios for Paris in 2040, aiming to explore potential urban developments at a mid-term horizon. Across all scenarios, we assumed that a certain proportion of each IRIS was subject to greening interventions, and applied our calibration factor to estimate the projected resulting increase in NDVI at the IRIS level. We only considered public greening interventions (by the municipality), excluding private greening initiatives.

The first scenario, S1, replaces removable parking spaces with green areas. For this, we retrieved the location and size of the majority of on-street payable car parking spaces, excluding 2 wheels, electric or priority status such as delivery and disabled parking spots, which represent 37.8% of all parking spots in the city. For all 20 districts of Paris, we then replaced a specific share of those parking areas with green spaces. These proportions, previously assessed by the French national institute of statistics and economic studies (INSEE), were based on the per-district share of residential on-street parking supply considered excessive, after deducting the surplus residential supply in facilities such as housing, concession and commercial parking (Atelier parisien d’urbanisme, 2019) (Supplementary Figure S3). The corresponding proportion of surface area represented by this transformation is represented on Supplementary Figure S4. Parking delineation data are publicly available (Direction de la Voirie et des Déplacements - Ville de Paris, 2025).

In the second scenario, S2, we assume that 20% of all streets per IRIS would be converted into UGS. Street network data were used to quantify, for each IRIS, the corresponding proportion of surface area represented by this transformation (Supplementary Figure S5). Street delineation data are publicly available (Direction des Espaces Verts et de l’Environnement - Ville de Paris, n.d.).

The third scenario, S3, is a best-case scenario. Here we simulated the implementation of green spaces sufficient to reach at least 15.41% municipal green space coverage within each IRIS. This target value corresponds to the 90^th^ percentile of green space coverage across IRIS, excluding those fully greened (Supplementary Figure S6).

### Health impact assessment

We conducted a health impact assessment of the change in NDVI on all-cause mortality in the city of Paris at the IRIS level. Demographic data were based on INSEE projections (INSEE, (National Institute of Statistics and Economic Studies), n.d.). To respect feasibility of implementation and the assessment of near-term co-benefit, we chose 2040 as an endpoint for our health impact assessment. We retrieved the age-specific population and mortality projections for France in 2040 and the population distribution at the IRIS level from 2019. To get the population size and mortality rate for each age in all IRIS for 2040, we applied the nationwide distribution of age-specific population size and mortality rates to the population of each IRIS. Based on a log-linear exposure-response relationship quantifying a reduction in all-cause mortality for an increased NDVI, we estimated the number of deaths prevented as:

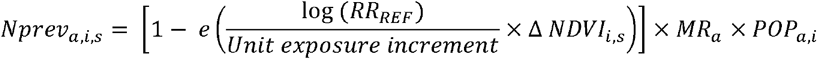

And:

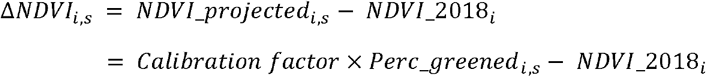

With:

*Nprev*_*a,i,s*_: Number of deaths prevented per age group *a*, in IRIS *i*, due to scenario *s*.

*RR*_*REF*_: Relative risk of all-cause mortality associated with the unit exposure increment, estimated from Cox proportional hazards models (Rojas-Rueda et al., 2019).

Δ *NDVI*_*i,s*_: Difference between projected and baseline NDVI, for each IRIS unit *i* and scenario *s*.

*Unit exposure increment* : Increase in NDVI associated with the *RR*_*REF*_.

*MR*_*a*_ : Mortality rate of age category *a*.

*POP*_*a,i*_ : Population size of age category *a* and for each IRIS unit *i*.

*NDVI*_2018_*i*_ : Baseline level of NDVI in 2018 in IRIS unit *i*.

*Calibration factor* : Cumulative estimated increase in NDVI (treatment effect) resulting from the greening of an entire IRIS (normalized dose).

*Perc_greened*_*i,s*_ : Projected proportion of IRIS unit *i* greened in scenario *s*.

*RR*_*REF*_ was estimated in a meta-analysis at 0.96 [95% CI: 0.94-0.97] for an increase of 0.1 in NDVI (Rojas-Rueda et al., 2019). The unit exposure increment represents the change in NDVI yielding a risk reduction of *RR*_*REF*_. Specifically, an increase of 0.1 in NDVI has been linked to a 4% reduction in all-cause mortality (Rojas-Rueda et al., 2019). We restricted our assessment to the population aged over 20 years to correspond to the cohorts included in the meta-analysis (Rojas-Rueda et al., 2019). In a second step, this allowed to derive life-years gained based on the projected life expectancy estimated by INSEE. Life-years gained were computed as the difference between life expectancy and age at the time of death.

All the estimates were presented alongside uncertainty intervals (UI), that we calculated using a Monte-Carlo simulation approach in order to propagate uncertainty in the outcome results. This included uncertainty arising from the estimation of the calibration factor linking the size of greening interventions to NDVI evolution, as well as the uncertainty in the exposure-response relationship linking NDVI and all-cause mortality. Specifically, 1,000 simulations were performed by independently drawing random values from log-normal distributions for each parameter. Final results were summarised as medians and 95% uncertainty intervals based on percentiles 2.5 and 97.5.

### Social deprivation

We present mortality impacts stratified by social deprivation level. This was done using the French deprivation index (FDep, 1997-2001) (Rey et al., 2009) and, in a supplementary analysis, using the European Deprivation Index (EDI, 2006) (Pornet et al., 2012), both available at the IRIS level. To analyse the distribution of mortality benefits according to those indexes, we categorized them into five quintiles, the first one representing the most favoured areas. We present the mortality benefits across deprivation levels using the preventable mortality fraction to account for the population density difference across IRIS and deprivation levels. Mortality rates and exposure-response functions are assumed uniform across deprivation levels.

The FDep index is defined as the first principal component obtained from a principal component analysis of four variables: the median income per consumption unit in households, the percentage of high school graduates in the population over 15 years of age, the percentage of manual workers in the active population, and the unemployment rate. This index was chosen because of its strong and consistent association with mortality in France, providing a reliable measure of spatial socioeconomic heterogeneity to address spatial health inequalities (Rey et al., 2009). The geographical distribution of these four variables are publicly available (INSERM, 2019).

### Cost-benefit analysis

For our economic assessment, we conducted a cost-benefit analysis on the 2025-2050 period. We estimated the implementation cost of each scenario following three hypotheses about different types of urban greening intervention (Table 1). For this, we relied on unit prices (Euros per m^2^) provided by an open-access dataset with quantitative estimates for urban soil restoration costs (Salin and Claron, 2025). This dataset is based on 56 semi-structured interviews and a survey of technical documentation. It is accompanied by a dedicated Excel spreadsheet that allows the construction of greening scenarios and the estimation of their median costs. These scenarios involve combining various techniques in the greening process, including preliminary studies, building demolition, unsealing, soil improvement, waste and excavated soil management, planting of vegetation, monitoring and maintenance. Supplementary text 2 provides more details on the greening intervention hypotheses. In **hypothesis A**, existing soils are improved after being unsealed without the addition of external topsoil, using ecological and soil engineering techniques such as decompaction, bio-inoculation and seeding or planting of herbaceous vegetation. In contrast, **hypothesis B** proposes a more conventional restoration process, albeit with less attention paid to preserving both urban and agricultural soils. This involves replacing the unsealed soil with a mixture of topsoil and compost and carrying out rapid revegetation by planting mature or semi-mature trees. As Scenario 3 makes no assumptions on the location of greening intervention, we extended hypothesis B by adding the demolition of building and occasional asbestos removal to set up **hypothesis C**. This corresponds to a logic of major transformation, typical of an urban park project. Costs related to blue space interventions were not included in any hypotheses. We equally shared the costs from 2025 to 2035 and added a 15 €/m^2^ maintenance cost each year from 2035 to 2050. The specific technical operations corresponding to hypotheses A, B and C are presented on Table S1.

**Table 1.**
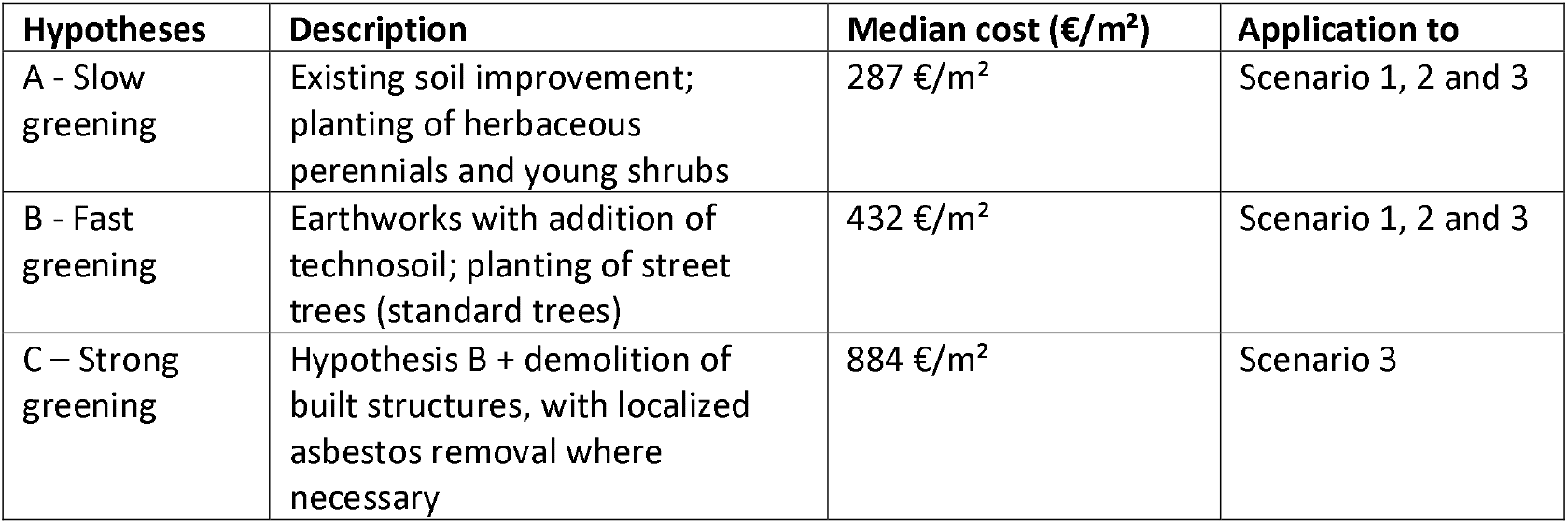
Description of our three different hypotheses for urban greening interventions, along with their median costs and associated scenarios in which we applied them.

| Hypotheses | Description | Median cost (€/m<sup>2</sup>) | Application to |
| --- | --- | --- | --- |
| A - Slow greening | Existing soil improvement; planting of herbaceous perennials and young shrubs | 287 €/m<sup>2</sup> | Scenario 1, 2 and 3 |
| B - Fast greening | Earthworks with addition of technosoil; planting of street trees (standard trees) | 432 €/m<sup>2</sup> | Scenario 1, 2 and 3 |
| C – Strong greening | Hypothesis B + demolition of built structures, with localized asbestos removal where necessary | 884 €/m<sup>2</sup> | Scenario 3 |

The benefits of each scenario were computed based on the estimated monetary value of life-years gained with the interventions. To estimate these health-related economics benefits, we applied the Value of a Statistical Life Year (VSLY) that is recommended in France for the socioeconomic evaluation of public investments (E.Quinet, 2013). The VSLY is a macro-economic indicator reflecting the monetary value society attributes to a marginal reduction in mortality risk. It was developed to support decision-makers in evaluating the cost-effectiveness of public health and environmental policies (Kniesner and Viscusi, 2019). In France, the VSLY is projected to reach €215,000 in 2050. The VSLY was multiplied by the number of life-years gained to estimate the monetised mortality benefits associated with reduced mortality. All monetised values were expressed in €_2025_. To account for the 5-year time period between implementation and NDVI evolution with our calibration factor (Table S2), we simulated a linear increase of NDVI between 2025 and 2040, and held it constant until 2050.

While the use of discount rates is a standard practice in economic evaluations of future costs and benefits, its application in the context of environmental changes (such as ecosystem degradation or restoration) or health evaluation issues is subject to ethical and technical controversy (Polasky and Dampha, 2021). In the main analysis, no discount rate was applied to mortality benefits and a 3% discount rate was applied to costs. This methodological choice reflects a commitment to intergenerational equity, recognising that decisions made in the present will have profound and lasting consequences for future generations. Moreover, the anticipated mortality and economic damages from climate change are expected to escalate significantly over time (IPCC (Intergovernmental Panel on Climate Change), 2023). Delaying climate adaptation policies now is likely to result in substantially higher costs—both economic and societal—in the future, it is therefore essential to value future benefits rather than diminish their importance through discounting (Stern, 2018). We applied a classical 3.2 % discount rate to both costs and benefits in a supplementary analysis to align with national recommendations (France Stratégie & Direction générale du Trésor, 2021).

### Sensitivity analysis

We conducted four sensitivity analysis to assess the robustness of our findings. First, we used the mean NDVI value of the six IRIS units identified as fully greened as a calibration factor (e.g., *Buttes-Chaumont, Bois de Vincennes, Bois de Boulogne*). As these areas represent the upper limit of vegetation cover in Paris, we assume that this value reflects the maximum reachable NDVI in Paris, corresponding to a ceiling value for the calibration factor. Second, to evaluate the influence of demographic projections, we repeated the analysis using a static demographic structure based on 2018 population data. Third, we investigated the spatial sensitivity of our exposure assessment by adding a 500-meter buffer zone around each IRIS unit, accounting for potential cross-border exposure. Finally, we applied a linear exposure-response relationship to assess the impact of the linearity assumption on our estimates.

## Results

### Calibrating and projecting NDVI evolution

In 2018, public green space coverage in Paris is around 23.8% (2500 Hectares), and the population-weighted mean NDVI was 0.240 (range 0.012 – 0.694). The increase in NDVI (Figure 1A) and projected resulting NDVI (Figure 1B) across IRIS are represented on Figure 1 for each scenario. In Scenarios 1 and 2, replacing surplus parking supply and 20% of street surfaces with green spaces would result in the greening of respectively 77 and 272 hectares, which represent 0.7% and 2.6% of total Paris area, respectively. Scenario 3, which targets a minimum green space coverage of 15.4% in each IRIS, requires the implementation of 917 additional hectares of UGS, representing 8.7% of the total city surface area.

**Figure 1.**
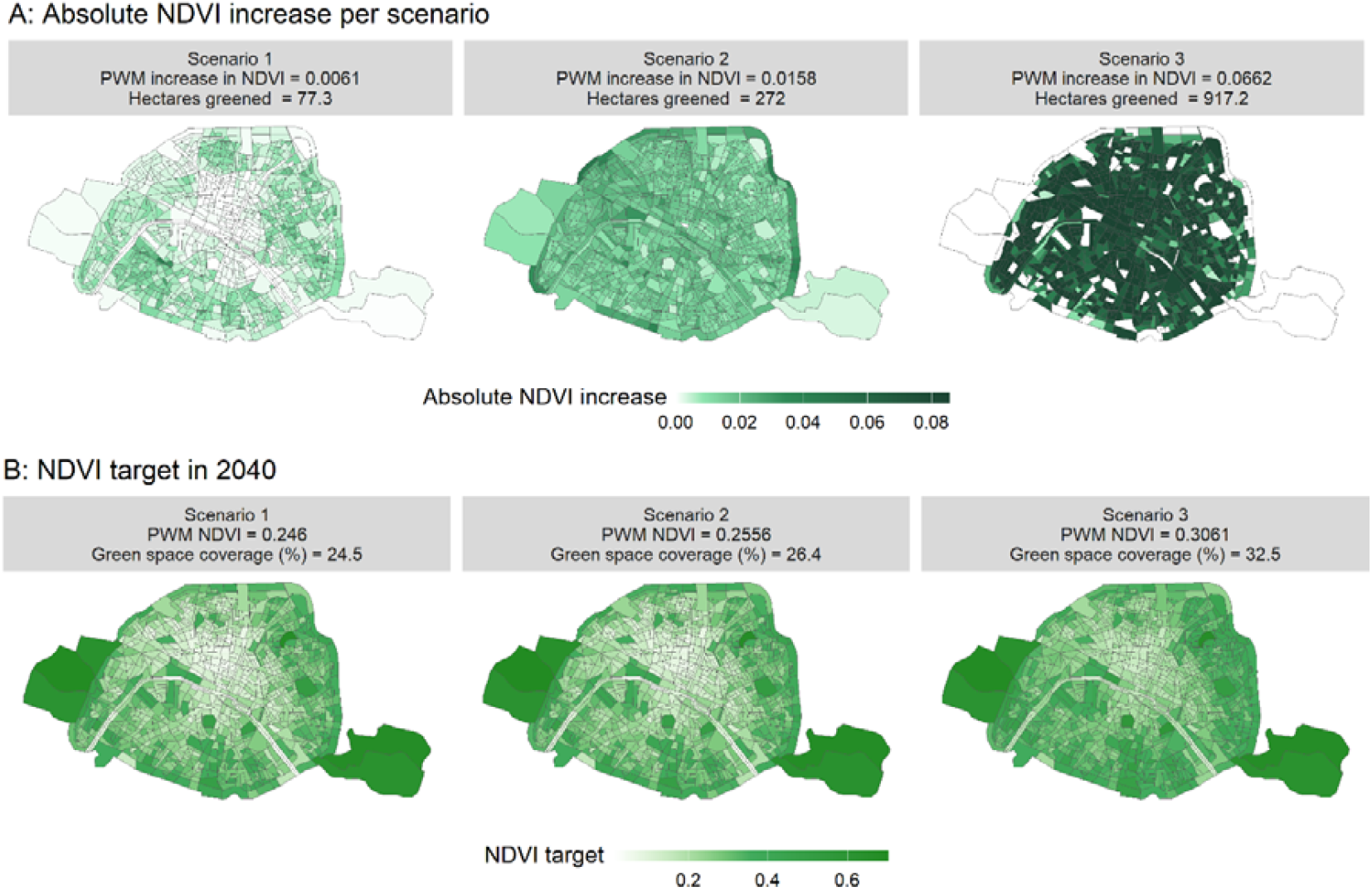
Absolute increase in NDVI (A) and projected resulting NDVI (B) across scenarios with associated population-weighted mean (PWM) value, hectares greened and final green space coverage (%). Scenario 1, 2 and 3 respectively correspond to replacing surplus parking, replacing 20% of street and reaching 15.4% of vegetation cover in each IRIS.

The average cumulative increase in NDVI in a fully greened IRIS was estimated at 0.56, 95% confidence interval [CI]: 0.40–0.71 (Supplementary Table S2). This means that a greening intervention of an entire (100%) IRIS would increase NDVI by 0.56, the greening of half (50%) an IRIS would increase NDVI by 0.28, and so on, as we computed a normalized effect allowing a dose-response interpretation. This corresponds to the average total effect of greening interventions based on observed NDVI changes following the 113 greening interventions >9001m^2^ implemented in Paris between 2001 and 2017 (Supplementary figure S1). The yearly effect of greening interventions consistently ranged between 0.08 and 0.14 across all post-treatment years, indicating a sustained increase in NDVI following interventions. The estimated average number of post-treatment periods contributing to the effect estimation is 5.4. This indicates that, across all treated IRIS units, outcome trajectories contributed to the estimation of treatment effects for, on average, five post-treatment time periods—in other words, the average timeframe needed for a greening intervention to have a normalized effect of 0.56 on the NDVI is 5.4 years.

### Quantitative health impacts assessment

Figure 2 shows the estimated number of deaths prevented under each scenario for 2040, along with the corresponding deaths prevented per hectare of UGS implemented. For 2040, scenarios 1 and 2 resulted in an estimated 29 (95% UI: 17–43) and 75 deaths prevented (95% UI: 43–112), respectively. Scenario 3, which simulates a minimum green space coverage of 15.4% within each IRIS units, leads to an estimated 312 deaths prevented (95% UI: 180–462) in 2040. These deaths prevented correspond to, respectively for scenarios 1, 2 and 3, 0.2% (95% UI: 0.1–0.3), 0.6% (95% UI: 0.4–0.9) and 2.6% (95% UI: 1.5–3.8) of mortality at the city level. Mortality benefits per unit area (influenced by population density) are relatively consistent across scenarios, with around 0.33 deaths prevented per hectare greened. As scenarios 1 and 2 can be jointly implemented, greening car-dedicated space could prevent around 100 deaths in 2040, representing 0.8% of the total mortality in Paris.

**Figure 2.**
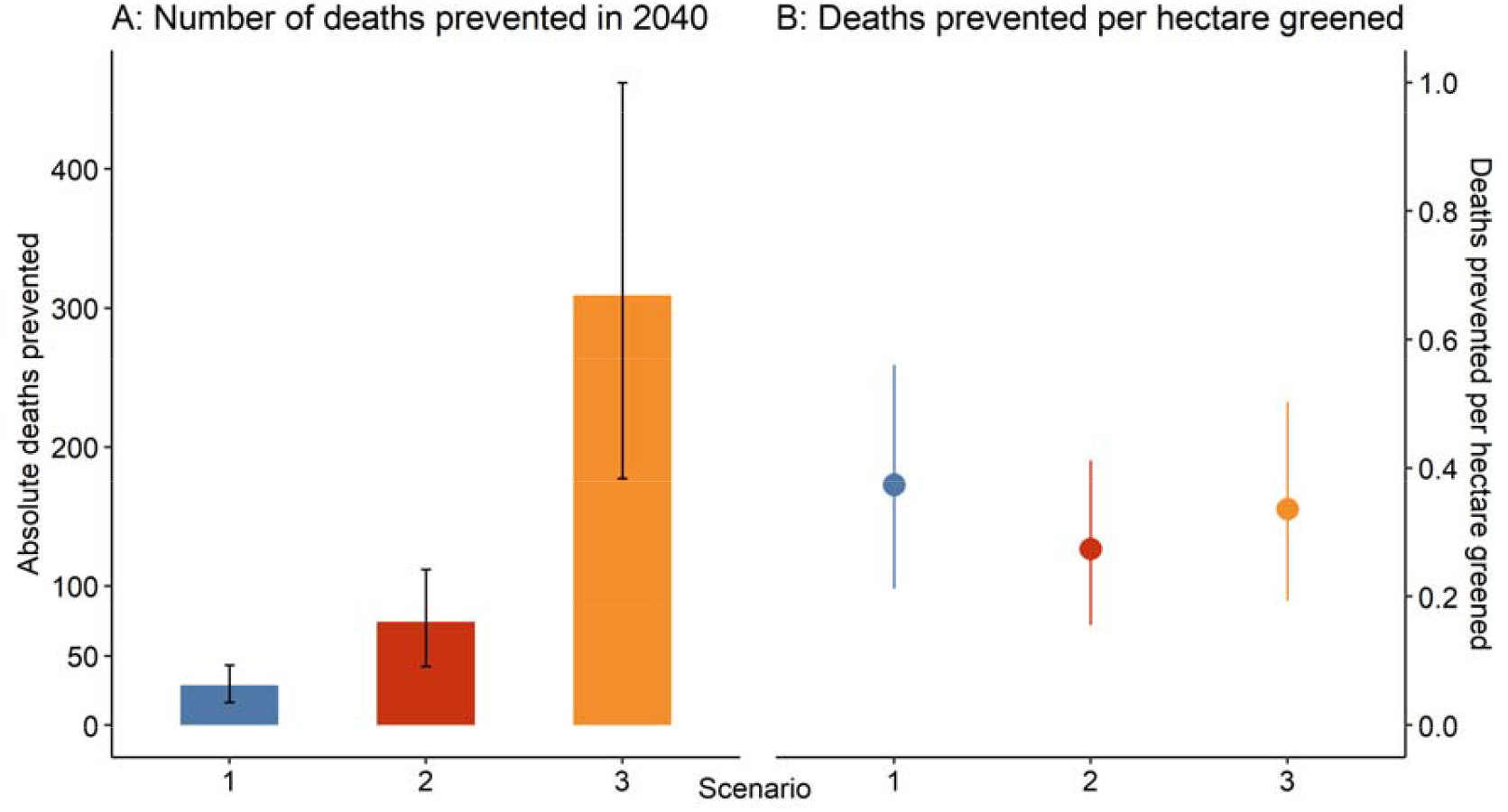
Number of deaths prevented in 2040 (A) and associated deaths prevented per hectare greened (B) across scenarios. Scenario 1, 2 and 3 respectively correspond to replacing surplus parking, replacing 20% of street and reaching 15.4% of vegetation cover in each IRIS.

All scenarios generated mortality benefits across all age groups, with deaths prevented particularly among older adults (≥70 years old), while younger age groups (20–50 years) contributed more substantially to life-years gained (Supplementary figure S7).

### Health impacts regarding deprivation index

Figure 3 presents the distribution of NDVI levels and estimated preventable mortality fractions across FDep quintiles for each scenario in 2040. The most favoured areas are concentrated in the center-western part of the city (Figure 3A). NDVI baseline levels in 2018 were similar across FDep score areas, with slightly higher values for the most deprived areas (FDep quintile = 5, Figure 3B). Under Scenarios 1 and 2, the preventable mortality fractions were stable across deprivation levels (Figure 3C). Scenario 3 yielded substantially higher mortality benefits, with preventable mortality fractions reaching around 2.5% in most quintiles. In this latter scenario, IRIS with higher deprivation exhibited marginally lower preventable mortality fractions, yet deprivation explained only a small proportion of the variance, with a Pearson correlation of r = 0.167 (95% CI [0.160; 0.174]) between mortality impacts and deprivation index, suggesting no meaningful differences in the mortality benefits across FDep scores. The same analysis was conducted with the EDI score; this index had a slightly different geographical distribution and associated NDVI levels but shows similar results regarding mortality benefits across deprivation scores for all scenarios (Supplementary figure S8).

**Figure 3.**
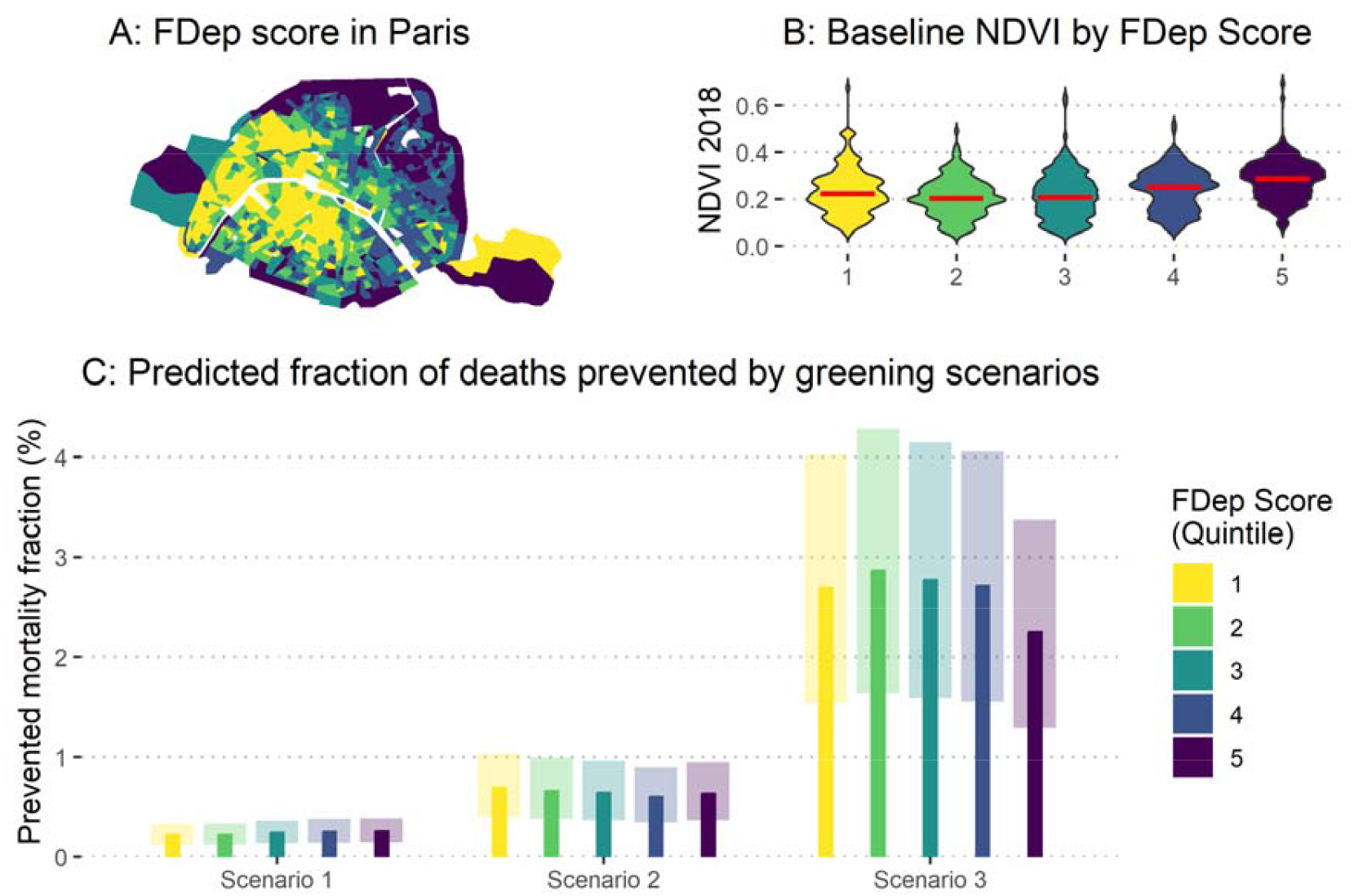
Geographical repartition of the FDep social deprivation score in Paris at the IRIS level (A), NDVI baseline exposure distribution in 2018 (B) and predicted fraction of deaths prevented by three greening scenarios (C) according to the FDep score. On all three panels, FDep is stratified into 5 quintiles, from less deprived (quintile 1, in yellow) to most deprived (quintile 5, in purple). On panel B, red bars represent median NDVI values. On panel C, scenario 1, 2 and 3 respectively correspond to replacing surplus parking, replacing 20% of street and reaching 15.4% of vegetation cover in each IRIS. The transparent portions depict the uncertainty intervals in the predicted mortality benefits due to uncertainties in the relative risk and calibration factor.

### Cost-benefit analysis

Figure 4 presents the predicted costs, benefits and cost-benefit ratio of the three considered greening scenarios. As the main costs associated with greening interventions were assumed to be incurred only once at the time of initial implementation, the cost-benefit ratio decreased over time (Figure 4A). Costs exceeded benefits at first but as the vegetation progressively increased, the benefits derived from mortality benefits rapidly outweighed intervention costs. In the long term, the benefits cumulated over 25 years were 1.4 to 3.7-fold higher than the cumulative costs, depending on the scenario and greening intervention hypotheses (Figure 4B). All three greening intervention hypotheses ended up cost-effective when cumulating economic impacts over time. Accounting for a discount rate of 3.2% on costs and benefits substantially reduced the absolute benefits but did not change the evolution of the ratio (Supplementary figure S9). Differences in the cost-benefit ratio across scenarios (following similar cost assumptions) were driven by population density and age group repartition across scenario. As for the deaths prevented by hectare greened (Figure 2B), here the most cost-effective scenario appears to be scenario 1.

**Figure 4.**
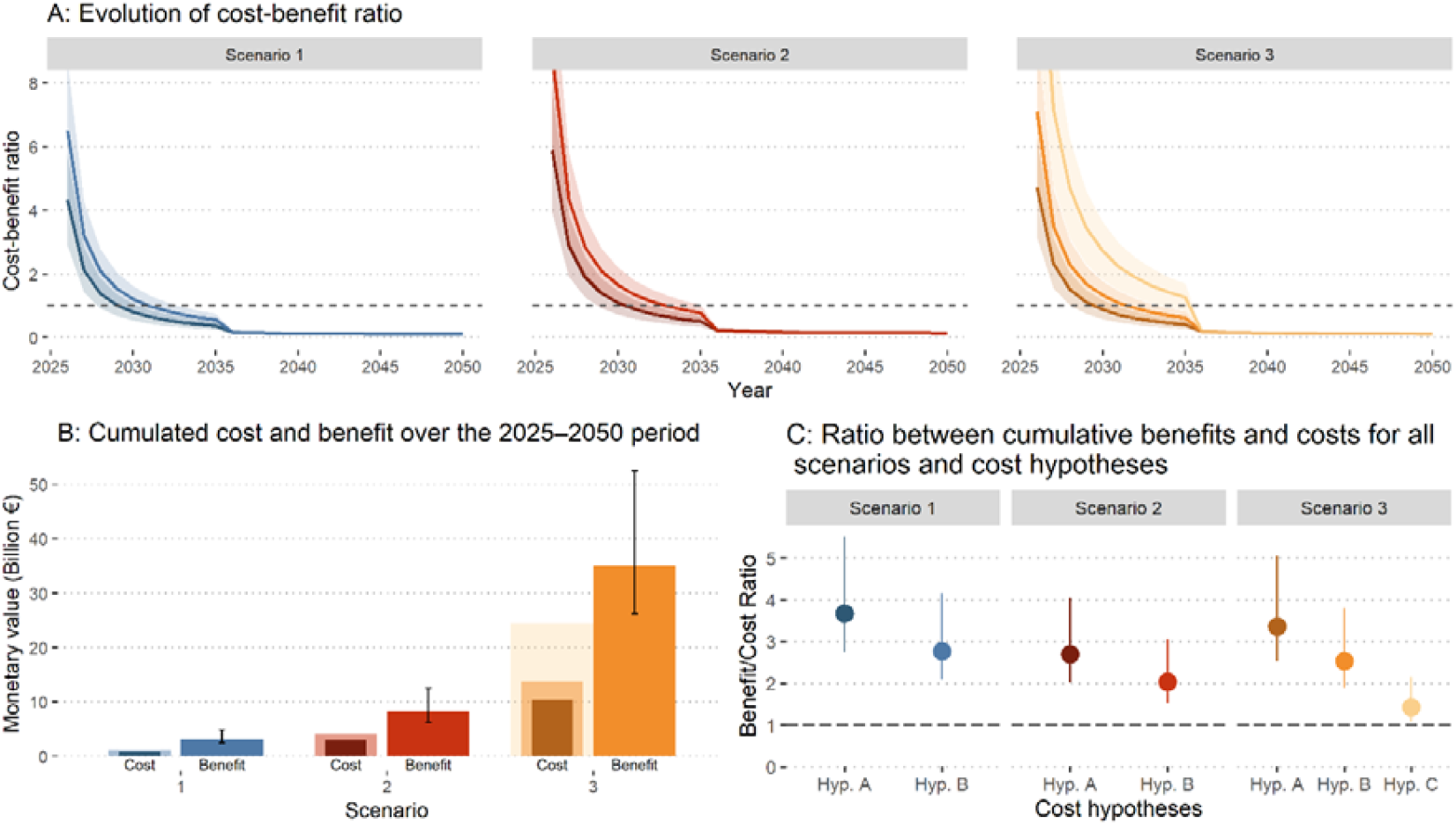
Evolution of cost-benefit ratio (A), cumulated costs and benefits (B) and ratio between cumulative benefits and cumulative costs (C) for each scenario and greening intervention hypotheses. Scenario 1, 2 and 3 respectively correspond to replacing surplus parking, replacing 20% of street and reaching 15.4% of vegetation cover in each IRIS. Greening intervention hypotheses are represented with different color intensity, ranging from dark (Hypothese A: Slow greening) to medium (Hypothese B: Fast greening) and light shades (Hypothese C: strong greening).

### Sensitivity analysis

The results of the sensitivity analysis are summarised in Table 2. The choice of an alternative “ceiling value” for our calibration factor had moderate effect in estimated benefits. In the second sensitivity analysis, using the population size, age-distribution and mortality rate of 2018 produced a minor reduction in our estimates as no major evolution of demographic dynamics are specifically projected for Paris. Adding a 500m buffer zone around IRIS units resulted in a small decrease in deaths prevented for 2040. Lastly, applying a linear exposure–response has minor impact on estimated benefits compared with a log-linear function. Overall, these findings suggest that our assessment is robust to variations in assumptions on calibration, population, spatial exposure and linearity in the exposure-response.

**Table 2.**
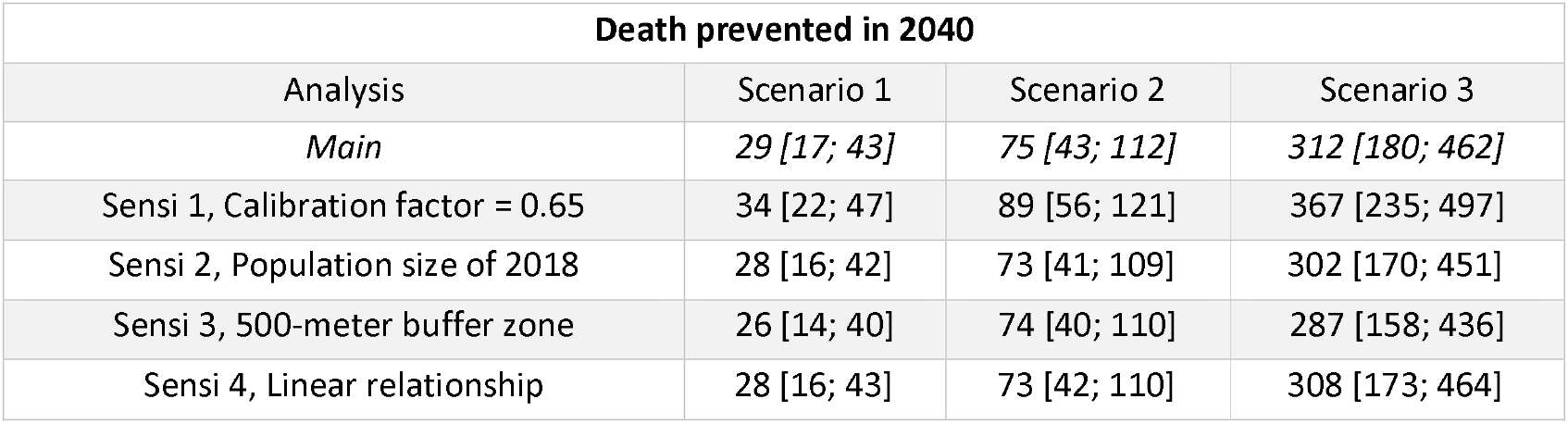
Main and sensitivity analyses of mortality benefits of greening interventions for all scenarios.

## Discussion

### Main results

Relying on an innovative approach combining a dynamic Difference-in-Differences model to compute exposure evolution in a health impact assessment, this study highlights substantial public health benefits of various urban greening strategies in a high-density context. As our framework quantifies health impacts per unit of greened surface area, we were able to conduct a cost-benefit analysis comparing the costs of greening interventions with the mortality-related benefits. We estimate that greening interventions reducing car-dedicated urban space (surplus parking or street spaces) could significantly improve health solely through exposure to green spaces, reducing all-cause mortality by up to 0.8% (Scenario 1 + 2). Reaching a minimum of 15% of green space coverage across all neighbourhoods could prevent around 300 deaths each year in Paris, representing approximately 3% of the city’s total mortality burden. Overall, one death can be averted yearly per every 3 hectares converted into greenspace, with consistent mortality benefits across socio-economic deprivation group. Finally, in all our scenarios, the monetised mortality benefits associated with exposure to UGS offset the implementation costs as soon as 2035, indicating net societal benefits in the long term. Qualitative results remained consistent across several sensitivity analysis.

### Literature comparison

As part of the C40 global network, the city of Paris is engaged in reaching 30-40% of green or permeable spaces and to provide an accessible green or blue space within a 15-minute walk to 70% of its residents by 2030 (C40 Cities Climate Leadership Group, n.d.). In 2024, the total municipal green space coverage was almost 2500 hectares, which represents ~24% of the city area. However, over 70% of this coverage is concentrated in peripheral wooded areas, while vegetation cover in the inner city remains limited at less than 6% (~600 ha). Although the city coverage is close to the C40 Urban Nature Accelerator commitment, the present spatial distribution of green infrastructure can partly restrain both climate and health benefits and equitable access across neighbourhoods. The implementation of the current city roadmap (which involves greening 145.5 additional hectares, representing 1.4% of the city surface) will by itself not be enough to reach the C40 cities target (Mairie de Paris, 2023). Further policies aimed at replacing car-dedicated space would accelerate progress toward these goals, while providing sustainable health benefits and a more biodiverse and resilient city. A health impact assessment study on the member cities of the C40 network estimated that uniformly increasing green space by 1% would prevent about 40 deaths (2 for 100 000 inhabitants) in Paris (Martin et al., 2025). This result is consistent with our estimations of 26 deaths (95% CI: 14-40) prevented with an increased coverage of 0.7% in the first scenario.

Using least squares quadratic regression, a health impact assessment of Philadelphia’s tree canopy estimated a 0.05 increase in NDVI for a 10% increase in tree canopy. Again, this result is consistent with our 0.07 increase in NDVI for an 8.7% increase in coverage in the third scenario (Kondo et al., 2020). Another health impact assessment study conducted across European cities estimated that achieving the WHO guidelines on accessible green space would correspond to a NDVI increase of 0.06 in Paris (greater city), reducing annual mortality by 4.9% (Barboza et al., 2021). Our third scenario increases the population-weighted mean NDVI by 0.06 and prevents around 3% of mortality. The relatively lower benefits observed in our analysis may be partly attributed to spatial heterogeneity captured in the NDVI values we used. The previous study relied on mean NDVI estimates calculated over the Greater Paris region, which includes areas with naturally higher greenness levels (forests or agricultural land) not representative of the denser urban centre. These higher values may mask the extent of greening needed within the city itself. Consequently, achieving the WHO NDVI recommendations in central Paris would likely require achieving a higher NDVI increase, with the associated health benefits reaching a large proportion of the population.

To quantify the relationship between greenspace implementation and NDVI evolution, our Difference-in-Differences approach offers a quasi-experimental framework with strong causal inference, when the parallel trend assumption is satisfied. This method is particularly useful for assessing the impact of specific local interventions, as it controls for unobserved time-invariant variables. It also allows to assess costs associated with greening scenarios, provided that local information on green space implementation costs is available. Alternatively, we could have used least squares quadratic regression, which offers a statistical description of the relationship between green areas and NDVI if the model is well fitted. This approach does not require a validity test to hold true, or greenspace planning data; however, it has less causal interpretation and is more sensitive to confounders. Directly using the mean NDVI value of fully greened local areas as a calibration factor, as tested in our first sensitivity analysis, is a simpler approach that could be explored in further studies to confirm its validity in various contexts.

### Implications of our findings

In France, 18% of the population does not visit UGS at least once a week, with a higher proportion of lowly educated individuals among UGS non-users (Łaszkiewicz et al., 2023). Considering environmental and social justice is essential when implementing UGS as they can have various effects on health inequities. Earlier studies suggested that mental and physical health benefits arising from UGS were stronger among the most deprived population (Belcher et al., 2024; Rigolon et al., 2021). However, greening interventions, if implemented without attention to local socio-economic dynamics, can foster gentrification and environmental inequalities (Kim and Wu, 2022; Oscilowicz, E, n.d.). Perceived safety also influences the use and health potential of green spaces by limiting its access, particularly for women, ethnic and gender minority, older adults or previously victimized individuals (Sreetheran and Van Den Bosch, 2014). To address these issues, greening interventions need to be associated with mechanisms to preserve housing affordability (rent control, community land trusts, inclusionary zoning) and participatory strategies involving local communities, ensuring that greening projects respond to the needs and voices of residents, especially those marginalized and vulnerable groups (Oscilowicz, E, n.d.). Good examples of such projects may be observed in Vienna and Amsterdam or, more specifically in France, in Nantes, where the city ensured green space accessibility to all residents within 300 meters, incorporated community participation in the urban planning process and required social housing quotas. Our study suggests that, with a constant repartition of deprivation levels in Paris until 2040, greening car-dedicated space would benefit equally to all deprivation level.

The configuration of green spaces (size, shape, and connectivity) modulates the magnitude of health benefits. Larger, more complex, and better-connected green areas are consistently associated with improved health (Wang et al., 2024). Given that parking lots and road spaces are well distributed throughout the city, our first and second scenarios would favour this connectivity thereby increasing associated health benefits. Redefining urban space with ambitious greening policies could also favour active mobility and provide substantial health co-benefits (Moutet et al., 2024). Furthermore, decisions around greening interventions can significantly influence costs, health benefits and environmental impact. For instance, opting for the planting of saplings combined with soil rehabilitation (greening intervention hypotheses A), rather than importing mature trees and external substrates (greening intervention hypotheses B), may delay ecological maturity but offers more affordable, locally adapted, and sustainable urban vegetation growth (Salin and Claron, 2025). Our analysis complements prior assessments of ecosystem services by adding monetised health benefits to the overall cost–effectiveness profile of nature-based solutions (González-García et al., 2025). Further studies exploring various greening scenarios specifically targeted at reducing health inequalities could use our Difference-in-Differences and cost-benefit framework to assess health impacts associated with different typology of policies (Benach et al., 2013).

Greenspaces may also have adverse effects on health that are important to consider in order to maximize health benefits. First, residential greenness has been associated with more severe allergy symptoms among pollen-sensitized adults (Stas et al., 2021). To support biodiversity while minimizing pollen exposure, UGS should prioritize selection of low-allergenic (avoiding birch trees, grasses and cypress) and insect-pollinated species, diverse plant communities and a balanced plant gender ratios (Stevanovic et al., 2025). In parallel, exposure to natural green spaces in early life can reduce the risk of allergic sensitization, potentially through beneficial effects on the gut microbiota and immune system development (Buchholz et al., 2023). Secondly, UGS can both mitigate and exacerbate the transmission of vector-borne diseases, depending on their structure, maintenance and local ecological context (Fournet et al., 2024). Thirdly, in dense urban areas such as Paris, air pollution may be locally affected in contrasting ways by urban trees. While UGS can improve air quality through deposit on leaf surfaces, they may also restrict NO_x_ dispersion and emit biogenic volatile organic compounds that contribute to ozone formation. To reduce these risks, planting of trees with large crowns on high-traffic streets should be avoided and tree species that emit few terpenes should be favoured in cities (Maison et al., 2024).

## Limitations

Our use of NDVI as a proxy for assessing the health impacts of green space exposure introduces several limitations. NDVI is a widely adopted measure that reflects vegetative greenness by capturing chlorophyll activity and has been widely used to explore associations between vegetation and mortality (Rojas-Rueda et al., 2019). However, first, NDVI indicates the presence of vegetation but does not account for its quality or functionality. Accessibility, availability and usability are critical parameters to achieve UGS benefits (WHO Regional Office for Europe, 2016). Integrating accessibility and biodiversity indexes could be a relevant and complementary approach to assess those policies, as well as monitoring ecologic quality and usage of UGS. Second, UGS encompass a diverse range of elements beyond vegetation (water, paved surfaces, recreational infrastructure…) that are not specifically captured by NDVI. This limitation is partially offset by our dynamic Difference-in-Difference framework that quantifies, within the local context of Paris, the increased NDVI associated with public UGS interventions, including vegetation and other infrastructures. Third, although not included in our analysis, private green spaces may contribute to mortality benefits and their distribution among socio-economic groups warrants further studies from an environmental equity perspective (De Vries et al., 2025). Fourth, as the demographic data (age distribution, size and mortality) were only available at the IRIS level, we used NDVI data at the same scale and therefore, did not capture potential heterogeneity in the exposure. This limitation motivated our 3^rd^ sensitivity analysis which shows that adding a 500-meter buffer zone around each IRIS does not significantly change the exposure assessment and therefore the associated health impacts.

Our analysis is also subject to temporal limitations, as our quantification of vegetation growth effects from urban greening interventions is constrained by the observation window (2000–2018); while the use of event-study Difference-in-Differences estimators allows to capture dynamic trends over time, full effects from the most recent greening interventions (post 2017) remain unobserved due to data availability. Integrating climatic projections and analysis of extreme weather events could also enhance NDVI projections and its association with human health.

Another potential limitation stems from our assumption of uniform mortality rates and exposure-response relationships across deprivation levels. Stratified mortality rate data were not publicly available at the IRIS scale and we did not identify specific exposure-response relationships, or any other effect modifier that would allow to specify the link between NDVI and all-cause mortality across deprivation levels. As mortality rates and population density are higher in most deprived area (Rey et al., 2009), and health benefits of greenspaces more pronounced in low socioeconomic status (Belcher et al., 2024; Wei et al., 2023), our estimates are likely underestimating health benefits, particularly in those areas. Future epidemiological research should investigate exposure-response functions stratified per socioeconomic status to better inform on greenspace-related health impacts, as it has been done for air pollution and mortality (Forastiere et al., 2007).

While we only considered health benefits in terms of prevented deaths and life-years gained, urban greening interventions improve mental health and well-being prior to measurable changes in mortality. However, while some mental health impacts can be estimated (Vidal Yañez et al., 2023), broader well-being effects remain difficult to quantify with current methodology. Spatial or demographic patterns in well-being tend to align with mortality, suggesting that conclusions similar to ours may be obtained when considering well-being effects (Martín-María et al., 2017). Urban vegetation also provides intergenerational health benefits such as reduced offspring asthma risk or increased masculine fertility, which are not included in longitudinal studies characterizing dose-response associations (Sinsamala et al., 2025; Zeng et al., 2025). These benefits would enhance the long-term benefits of UGS implementation. Further health benefits associated with active transportation, air quality or reduced noise can also be expected with greening interventions (Markevych et al., 2017). Lastly, in future years, with increasing temperatures, urban greening interventions are likely to provide a higher benefit on heat-related mortality (Wu et al., 2025).

## Conclusion

To our knowledge, this is the first study to use spatially explicit scenarios at a small scale to assess the mortality benefits of greening interventions. Using a dynamic over time Difference-in-Difference approach, we developed a framework based on a tangible urban design parameter: the surface area converted to green space. This framework can be adapted to other cities or types of intervention (e.g., cycling infrastructure, air pollution reduction measures or sustainable diet promotion) to estimate their health benefits and potentially their distribution across levels of social deprivation as well as their cost-benefit ratio.

Our results suggest that in Paris, repurposing car-dedicated urban space such as parking and street into green spaces would provide substantial mortality benefits while reinforcing the city’s climate mitigation and adaptation efforts. Approximately one death could be prevented annually for every three hectares of green space created. Establishing more ambitious targets for vegetation cover would further enhance these health co-benefits. Such interventions do not exacerbate health inequalities by themselves. Moreover, the associated monetised mortality benefits are expected to exceed the implementation costs, reinforcing the societal value of urban greening policies.

## Data Availability

All data produced in the present work are contained in the manuscript

## DECLARATIONS

### Availability of data and materials

All codes, analysis, extraction and quality grid are available in the following GitHub repository: https://github.com/LeoMoutet/eqis_evu_paris.

### Competing interests

The authors declare that they have no competing interests.

### Funding

This project did not receive any specific funding.

### Authors’ contributions

LM, LT and KJ designed the study. LM conducted data analysis. MP, JL and LA provided exposure data. KB contributed to local data acquisition and interpretation. MAB provided input regarding the Difference-in-Differences framework. LA contributed to analysis regarding social deprivation. CC constructed costs hypotheses and provided inputs for the economic assessment. LM wrote the original draft with input from LT and KJ. All authors contributed to critical feedbacks and approved the final manuscript.

## Notes

### Competing Interest Statement

The authors have declared no competing interest.

### Funding Statement

This study did not receive any funding

### Summary of Updates

Improving text, motivation and added value

