## Supplementary materials for "Replacing cars with green spaces: an assessment of the mortality benefits in Paris"

### Supplementary text 1: Details on the calibration

Here we use the ‘did_multiplegt_dyn’ package to estimate the causal effect of a treatment (greening intervention) on an outcome (NDVI evolution). This method, fully documented on Github (<https://github.com/chaisemartinPackages/did_multiplegt_dyn>) and formally developed in "Difference-in-Differences Estimators of Intertemporal Treatment Effects" by de Chaisemartin and D'Haultfœuille (2024), extends the Difference-in-Differences (DiD) framework to take into account time-varying and non-binary time-varying treatments (De Chaisemartin and D’Haultfœuille, 2024).

The did_multiplegt_dyn estimator is designed for group-level panel data—such as administrative units (IRIS) observed over multiple time periods—and accounts for both immediate and lagged treatment effects. It accommodates heterogeneous treatment timing across groups and allows treatment intensity to vary continuously over time.

To evaluate the cumulative impact of UGS interventions, the estimator implements a dynamic DiD design. For each treated unit and for each horizon 𝑙 = 1,2,…,𝑛, the change in NDVI relative to the pre-treatment baseline is compared to the corresponding change among control groups that have not yet been treated at that horizon.

These differences are then aggregated across time to estimate the average cumulative treatment effect over time. These group- and horizon-specific DiD contrasts are then aggregated to estimate average dynamic treatment effects. Aggregation weights reflect both:

1. Temporal coverage: The number of treated units observed at each future period.
2. Treatment intensity: The effects are further weighted by the size of the treatment, enabling a dose–response interpretation of the intervention’s impact. This dose was defined as the proportion of land area affected by the greening intervention within each IRIS unit.

The estimator relies on two key identifying assumptions: (i) a parallel trends assumption conditional on fixed effects whereby untreated potential outcomes would have followed similar trends across groups in the absence of treatment, and (ii) a no-anticipation assumption, implying that future treatment does not affect outcomes prior to implementation. Both assumptions are assessed empirically using pre-treatment placebo estimates (Figure S2). To assess the temporal dynamics of the intervention, we estimated post-treatment effects over a 14-year horizon. This yielded a series of event-study coefficients capturing the evolution of greenness (NDVI) following the implementation of urban greening interventions.

We apply a normalization procedure to express estimated effects per-unit treatment dose. Specifically, each event-study coefficient is divided by by the average incremental change in treatment intensity at the corresponding horizon (i.e., percentage of greened area), allowing for a dose–response interpretation of the results. In this specification, each coefficient can be interpreted as the average effect on NDVI of a one-unit increase in treatment exposure, conditional on time since treatment onset.

**Key Difference-in-Differences parameters:**

Treatment variable: Proportion of surface area converted into green space.

Panel structure: 992 IRIS units with annual NDVI observations from 2000 to 2018.

Treatment timing: Year of green space implementation.

Fixed effect structure: IRIS and year fixed effects.

Uncertainty estimation: IRIS-level clustered standard errors producing 95% confidence intervals.

**Difference-in-Differences estimator:**

$DID_{g,l}$ measures the causal effect of a treatment experienced by group g, evaluated $l$ periods after the change. The group- and horizon-specific DiD estimator is defined as follows (Equation 3 in de Chaisemartin and D’Haultfœuille, 2024):

$$DID_{g,l}=\left( Y_{g,F_{g}-1+l}-Y_{g,F_{g}-1} \right)-\frac{1}{N_{F_{g}-1+l}^{g}}\sum_{g^{'}:D_{g^{'},1}=D_{g,1},F_{g'}>F_{g}-1+l} (Y_{g^{'},F_{g}-1+l}-Y_{g^{'},F_{g}-1})$$

With:

$g$ : IRIS unit experiencing a treatment change.

$F_{g}$ : The **first year in which group** $g$ **changes its treatment (greening intervention).**

$l$ : The event-time horizon, $l$ ∈ {1, …, 14}.

$Y_{g,F_{g}-1+l}$ : The outcome (NDVI) of group $g, l$ periods after the treatment change.

$Y_{g,F_{g}-1}$ : The outcome at the year before the treatment change.

$N_{F_{g}-1+l}^{g}$ : Number of control (not-yet-treated) groups $g^{'}$, satisfying:

(i): $D_{g^{'},1}=D_{g,1}$: the same initial treatment level as group

(ii) : $F_{g'}>F_{g}-1+l$ : remain untreated at horizon $l$

$Y_{g^{'},F_{g}-1+l}-Y_{g^{'},F_{g}-1}$ : The evolution of outcome for each control group over the period $g$.

**Normalized aggregated Difference-in-Differences:**

Normalized event-study effect, aggregating group-specific normalized DiD estimates (Equation 15 in de Chaisemartin and D’Haultfœuille, 2024)

$${DID}_{l}^{n}= \frac{1}{N_{l}} \sum_{g:F_{g}-1+l \leq T_{g}} \frac{\left| \delta_{g,l}^{D} \right|}{\delta_{l}^{D}} \frac{DID_{g,l}}{\delta_{g,l}^{D}}$$

With:

$N_{l}$ : The number of groups that can contribute to horizon $l$ (i.e., groups for which the counterfactual control set is non-empty, $g:F_{g}-1+l \leq T_{g}$).

$\delta_{g,l}^{D}$ : Cumulative treatment dose, defined as the total amount of treatment received by group $g$ from the time of adoption to horizon $l$.

$\delta_{l}^{D}$ : Average cumulative treatment dose across all groups contributing to horizon $l$.

$\frac{DID_{g,l}}{\delta_{g,l}^{D}}$ : Group- and horizon-specific DiD effect per unit of cumulative treatment dose.

This normalization ensures a dose–response interpretation of the dynamic treatment effects and prevents groups with very small treatment intensities from exerting disproportionate influence on the aggregated estimator.

### Supplementary text 2: Cost estimates for urban greening intervention in Paris

This appendix establishes indicative cost ranges for greening public space in Paris and thereby justifies the financial assumptions adopted in our cost-benefit analysis.

1. Context and overview:

Since 2020 the city of Paris has launched urban-greening programmes, notably in the form of school playground conversions (“oasis playgrounds”) and “school streets” (“rues aux écoles”, when adjacent to a school). These projects typically strip impervious surfaces, introduce vegetation (planting beds, trees, flowering meadows) and add ancillary street furniture. The costs of these operations are high and vary considerably with the scale of works and techniques applied. (Salin et al., 2025) estimate the average cost of revegetating previously sealed ground in France at around €200-300/m² (typically the case of an asphalt street or courtyard to be desealed). However, data on actual developments of “school streets”, “oasis courtyards” or vegetated squares in Paris or comparable cities reveal significantly higher costs, due to two main factors:

**1/** **Urban density**. The intricate urban fabric of Paris, coupled with its sub-surface utility infrastructure and the constraints imposed by its heritage drive costs up.

**2/** **Comprehensive upgrades**. Available estimates for these operations include landscaping interventions and street furniture (tree grates, benches, planters, signage, etc.) – items of expenditure that are not taken into account in Salin's study, which focuses on interventions on the soil. A more realistic range for Parisian projects is therefore **€400–600 /m²**.

2. Observed orders of magnitude:

**Aggregate costs**

Between 2018 and 2022, Paris invested € 37 million in 94 oasis playgrounds, treating roughly 9 ha and unsealing 33 000 m² (4 % of the surface area treated)(Cour des comptes, 2023). In relation to the total surface area, this gives an average overall unit cost of **€41/m²**, but the figure masks heterogeneity (some sites received only light treatment) and works not aimed at greening. The conclusion of France's supreme audit institution (« Cour des comptes ») report on urban greening in Paris is worth noting: *Like any climate-adaptation policy, urban greening cannot be fully implemented if the primary focus is on its cost, without due consideration of the benefits that can be realised, and without the implementation of financial incentives (taxation, innovative financing, carbon tax). Aware of these issues, the City of Paris intends to undertake a study to quantify the benefits and avoided costs stemming from greening. (2023: 71)*

**Unit cost by project type**

For oasis playgrounds, CAUE 75 (2022) reports € 300–600 m², with potential savings expected from material reuse and community participation, although maintenance costs remain poorly documented (CAUE, 2022). For school streets, Paris communicated indicative budgets of € 300–800 000 per street during the 2022 participatory vote (Baverel, P., 2022); the lower figure was evidently political, as the Paris Climate Agency’s resource centre quotes € 700–800 000 for a fully vegetated street (Adaptaville. Agence parisienne du climat, 2025). Respire (2025) details component prices: planting beds, tree pits and wall greening cost € 180–550 (or € 2–5 k per planting unit), while a full street greening scheme reaches € 300–500 000 in Lyon and about € 800 000 in Paris. Although square-metre data are scarce, two internal memoranda of the City’s Directorate of Roads and Transport (City of Paris, Direction de la voirie et des déplacements, 2024, 2023) cite € 200–300 m² for pavement reconfiguration and € 500–600 m² for planting strips.

3. Ranges adopted in this study:

Synthesising the above, we retain unit costs of € 300–600 m² for oasis-type playgrounds and € 400–800 m² for school-street greening, and, for generic urban greening, a median order of €400-600 m². Expenditure in both cases generally comprises four items: roadworks (demolition and de-sealing, 30–50 % of the budget); landscaping (soil preparation, pedological engineering, topsoil, planting); street furniture (benches, play equipment, fencing); and long-term management, monitoring and maintenance, for which reliable figures remain limited. Cost variance is driven chiefly by the intervention scale (full versus partial pedestrianisation), the proportion of surface unsealed and vegetated, technical constraints (utilities, historic-building regulations, contamination) and the inclusion of additional equipment (e.g. fountains).

4. Building costs hypotheses:

We construct three urban-greening hypotheses and estimate their costs. The latter are drawn from the open-access urban-soil restoration database costs compiled by (Salin and Claron, 2025), on the basis of 56 semi-structured interviews and a review of technical documentation. The dataset is supplied with an Excel-based “scenario builder” that combines greening techniques and returns median cost estimates. The techniques considered include preliminary studies, demolition and pavement removal, soil improvement, waste and excavated-soil management, planting of herbaceous or woody species, and subsequent monitoring and maintenance.

Scenarios A and B rehabilitate sealed surfaces but diverge in soil treatment. Scenario A enhances the existing substrate through ecological engineering, whereas Scenario B undertakes extensive earthworks, installs technosoil, and plants mature trees to accelerate canopy formation. Scenario C adds the demolition of built structures—including, where necessary, asbestos abatement—and thus resembles the sweeping transformation typical of a new urban park.

### Figure S1: Map of the 113 green space interventions >900 m^2^ in Paris from 2001 to 2017

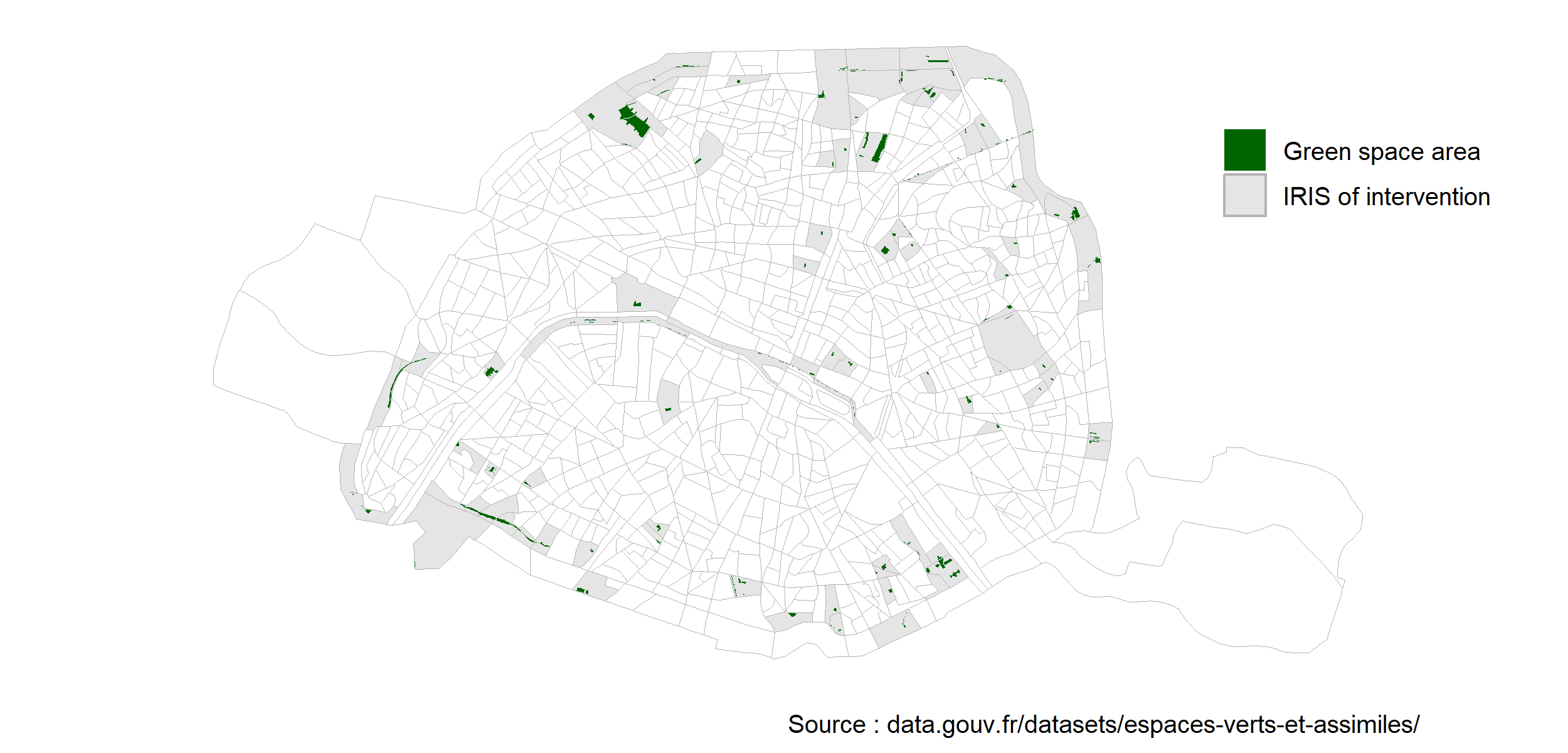

To build our calibration factor, as the NDVI is derived from a 30m resolution and computed for each IRIS as the mean summer value, and some interventions could be very small (<10 m^2^) we only included interventions >900 m^2^ for sensitivity (as 30m X 30m = 900 m^2^) representing the surface of the resolution.

### Figure S2: Effect of normalized greening interventions on NDVI in Paris using “did_multiplegt_dyn” on 2001-2017 period (output from ‘did_multiplegt_dyn’ package).

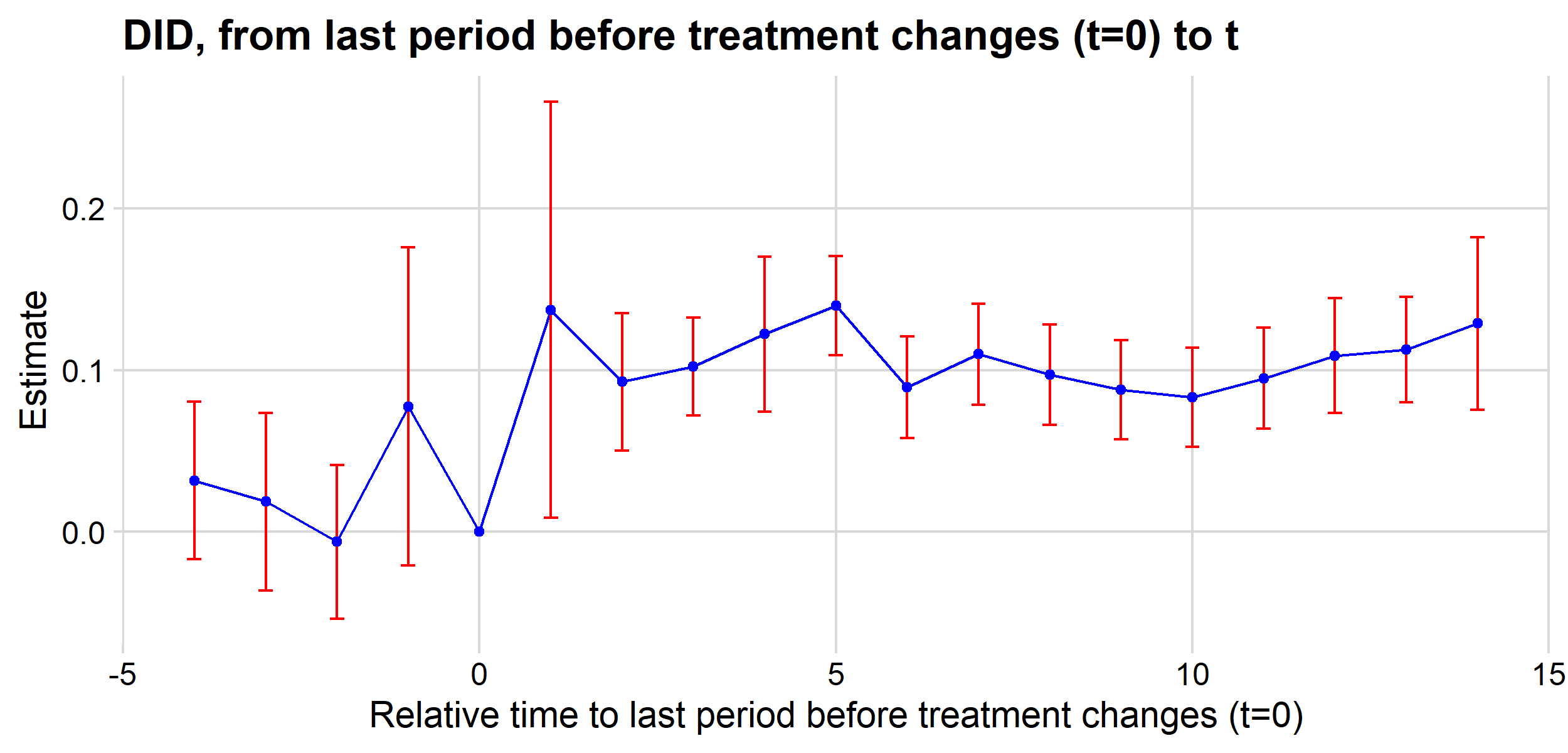

Test of joint nullity of the placebos : p-value = 0.4903.

P-value > 0.05, indicating no significant deviation from the main assumptions (« parallel trends » and « no anticipation ») thereby supporting the causal interpretation of the estimated effects.

### Figure S3: Scenario 1 – Proportion of surplus parking space per district

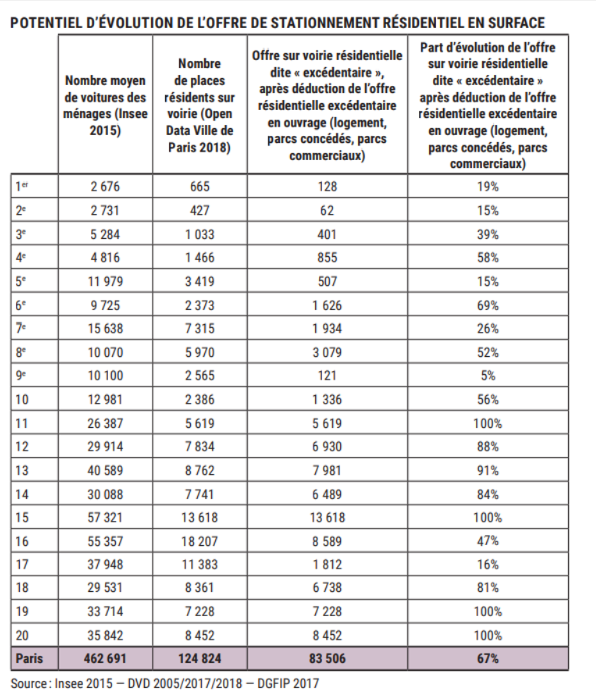

*English translation :*

**POTENTIAL FOR THE EVOLUTION OF THE SUPPLY OF SURFACE RESIDENTIAL PARKING**

Residential on-street parking supply considered « surplus », after deducting the surplus residential supply in other types of facility (housing, concession parks, commercial parks)

Share of the change Residential on-street parking supply considered « surplus », after deducting the surplus residential supply in other types of facility (housing, concession parks, commercial parks).

Average number of cars per household (Insee 2015)

Number of on-street resident parking spaces (Open Data Villes de Paris 2018

### Figure S4: Scenario 1 - Repartition of parking space (A), percentage of IRIS area occupied by a parking space (B), surplus parking space per district (C) and percentage of IRIS area occupied by surplus parking spaces (D)

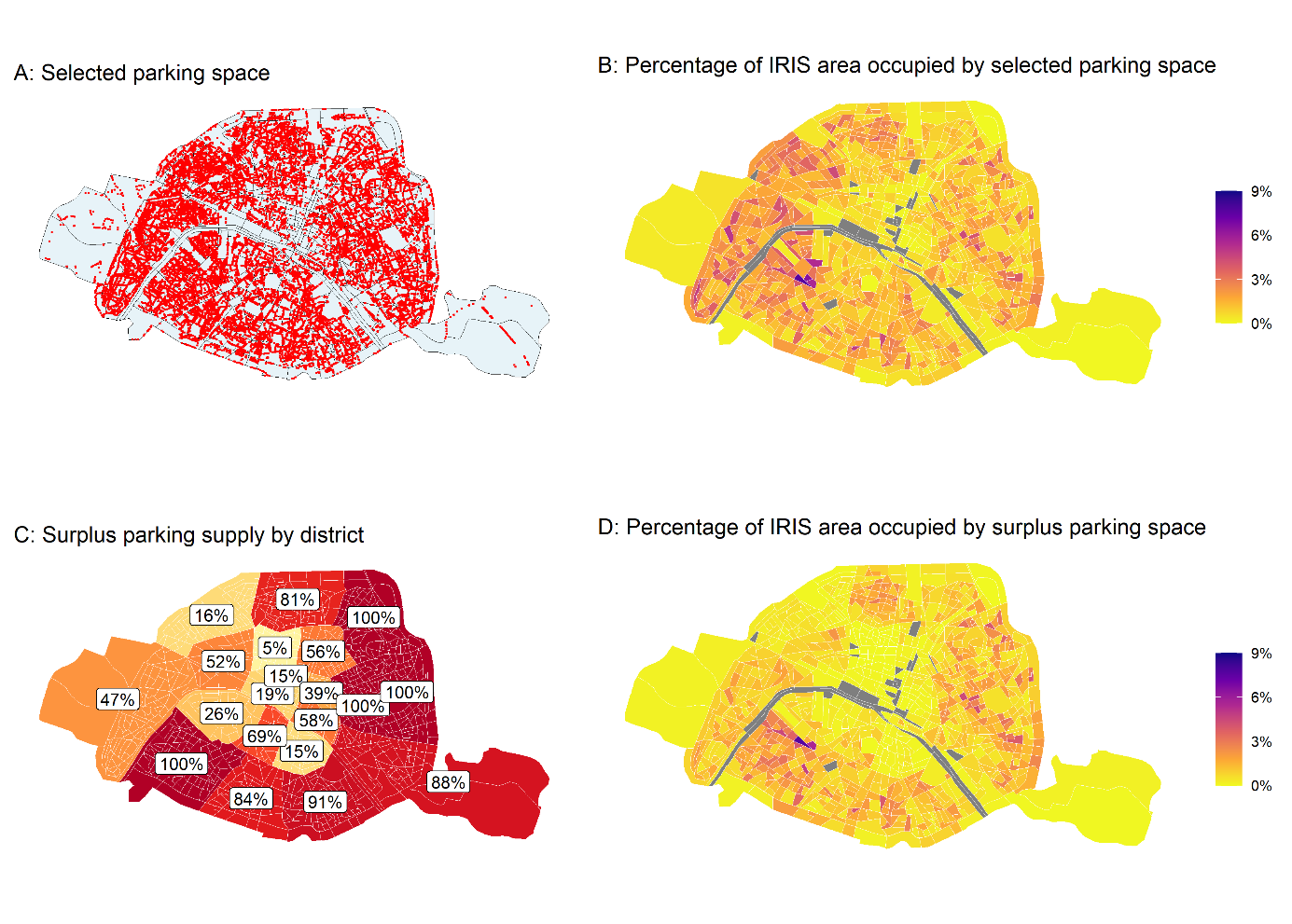

### Figure S5: Scenario 2 - Repartition of road space (A) and percent of IRIS surface taken by a 20% of road per IRIS (B)

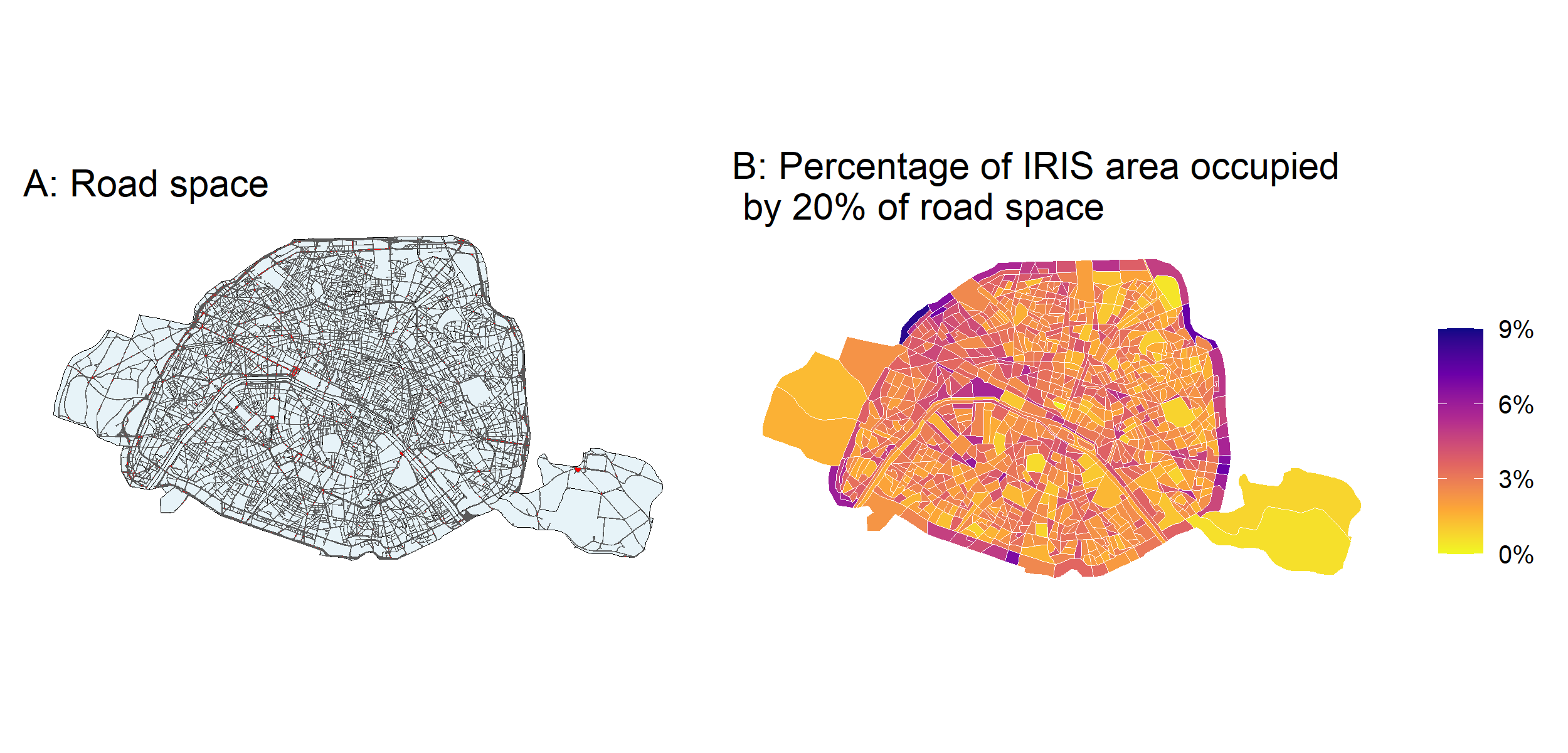

### Figure S6: Scenario 3 – Cut-off to reach green space coverage of the top 90th percentile of IRIS (excluding fully greened IRIS)

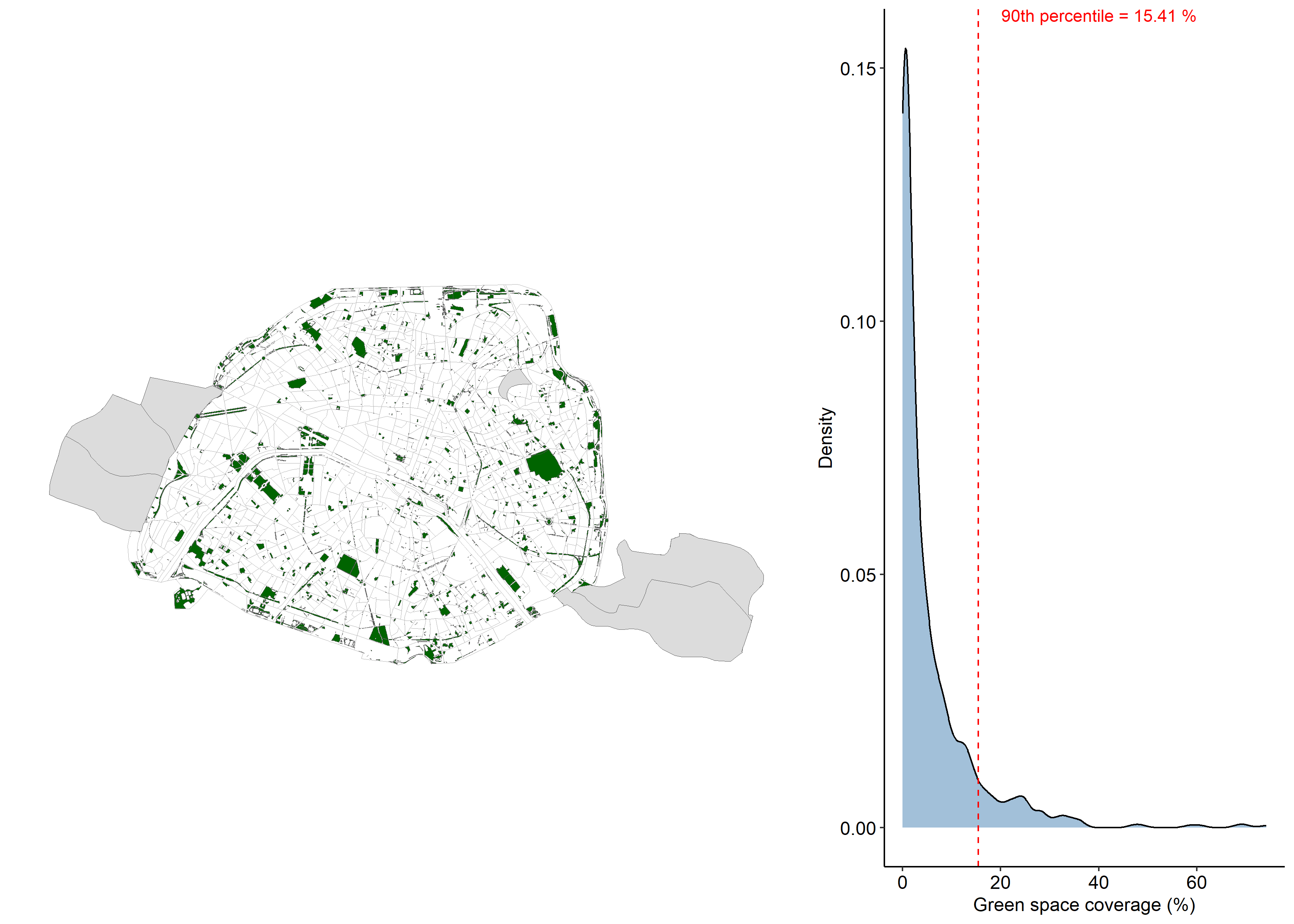

*Six IRIS (Vincennes, Boulogne and Buttes-Chaumont) are excluded as they are areas with no population, only composed of parcs, and their NDVI significantly differs from others.

In 2018 in Paris, total green space coverage was 2500 hectares, representing 23.7% of the city surface (603 hectares representing 5.7% when excluding those 6 IRIS).

### Figure S7: Distribution of deaths prevented and life-years gained by age group across all scenarios.

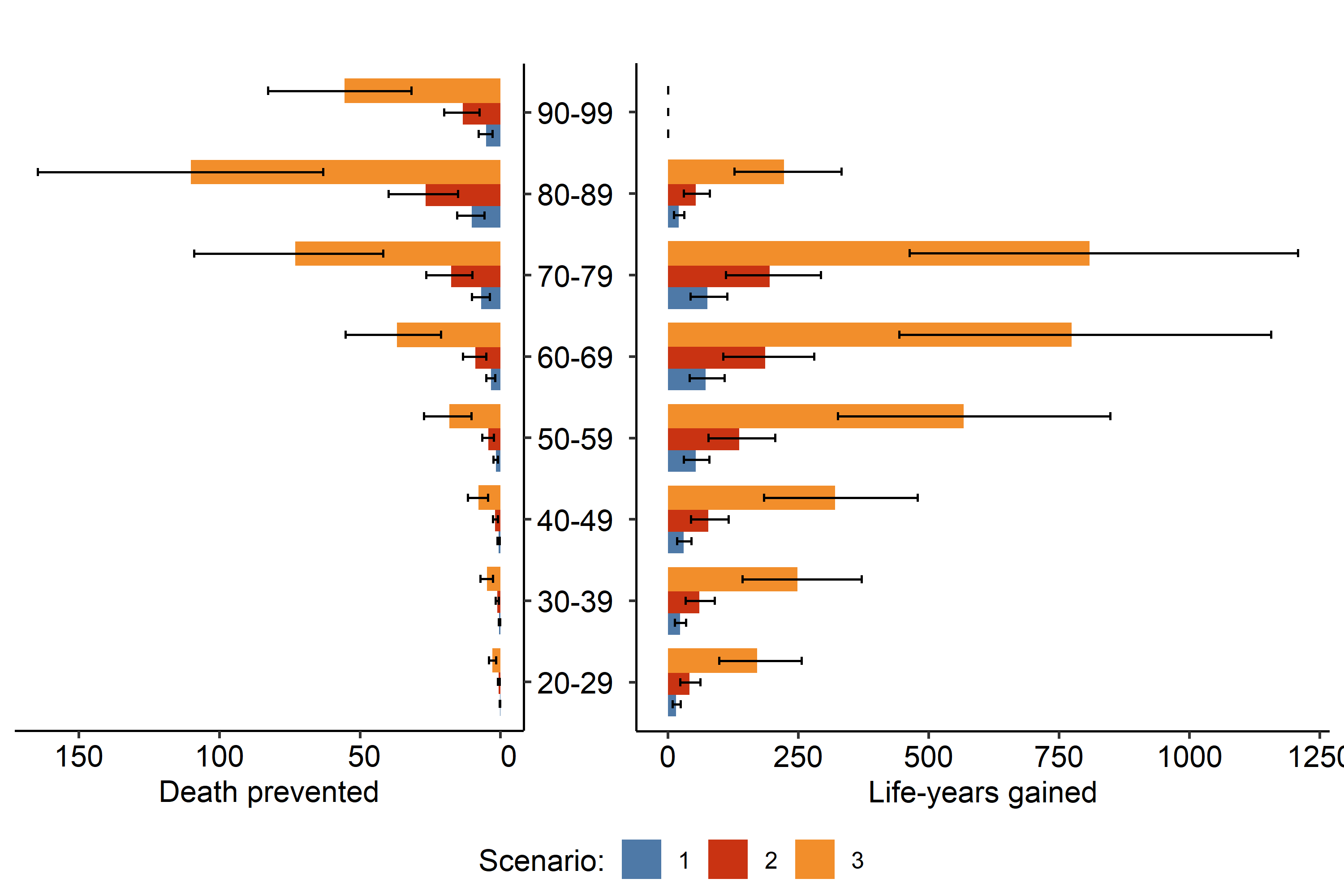

### Figure S8: Geographical repartition of the EDI deprivation score in Paris at the IRIS level (A), NDVI baseline exposure distribution in 2018 (B) and predicted fraction of deaths prevented by three greening scenarios (C) according to the EDI score.

On all three panels, FDep is stratified into 5 quintiles, from less deprived (quintile 1, in yellow) to most deprived (quintile 5, in purple). On panel B, red bars represent median NDVI values. On panel C, scenario 1, 2 and 3 respectively correspond to replacing surplus parking, replacing 20% of street and reaching 15.4% of vegetation cover in each IRIS. The transparent portions depict the uncertainty intervals in the predicted mortality benefits due to uncertainties in the relative risk and calibration factor.

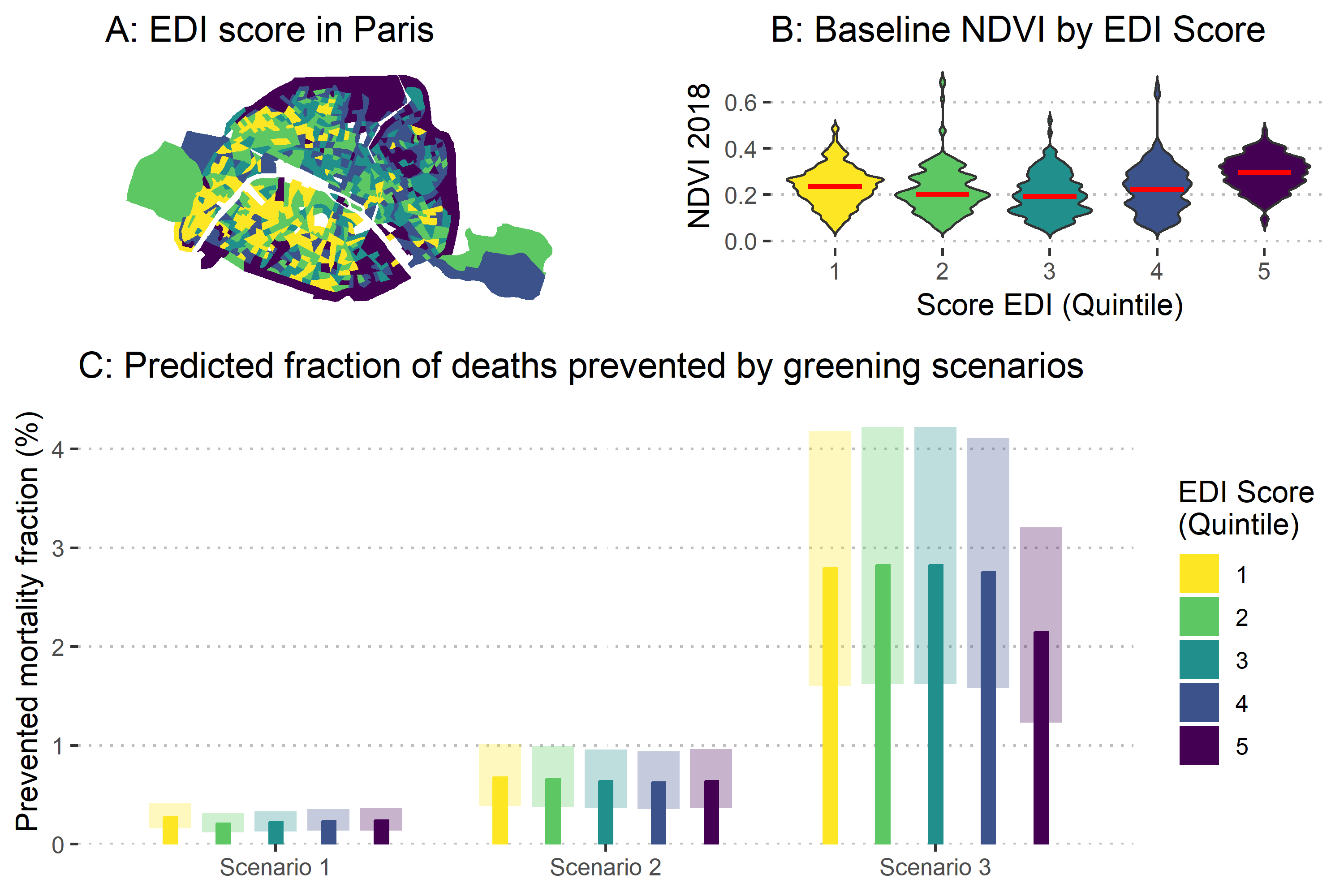

**Sources :** MapInMed, Université de Caen Normandie: European Deprivation Index (2017).

<https://storymaps.arcgis.com/stories/59026182c4b547c999a7048ec22c7498>

The EDI is a composite index designed to measure relative deprivation across European countries in a way that reflects individual experiences of social disadvantage. The French version of the EDI is based on a weighted combination of ten variables drawn from both individual-level data (EU-SILC survey) and aggregated census data. These variables are grouped into two core dimensions: material deprivation (such as non-homeownership, absence of a car, overcrowded housing, and large household size) and social deprivation (including unemployment, low-skilled occupations, low educational attainment, foreign nationality, single-parent households, and living alone). It incorporates both objective indicators of poverty and the subjective experience of unmet basic needs, such as being unable to handle an unexpected essential expense. The EDI offers a nuanced and standardized tool to monitor social inequalities in health, guide public policy, and support targeted interventions where they are most needed.

### Figure S9: Evolution of cost-benefit ratio (A), cumulated costs and benefits (B) and ratio between cumulative benefits and cumulative costs (C) for each scenario and greening intervention hypotheses.

Scenario 1, 2 and 3 respectively correspond to replacing surplus parking, replacing 20% of street and reaching 15.4% of vegetation cover in each IRIS. Costs hypotheses are represented with different color intensity, ranging from dark (Hypothese A: Slow greening) to medium (Hypothese B: Fast greening) and light shades (Hypothese C: strong greening).

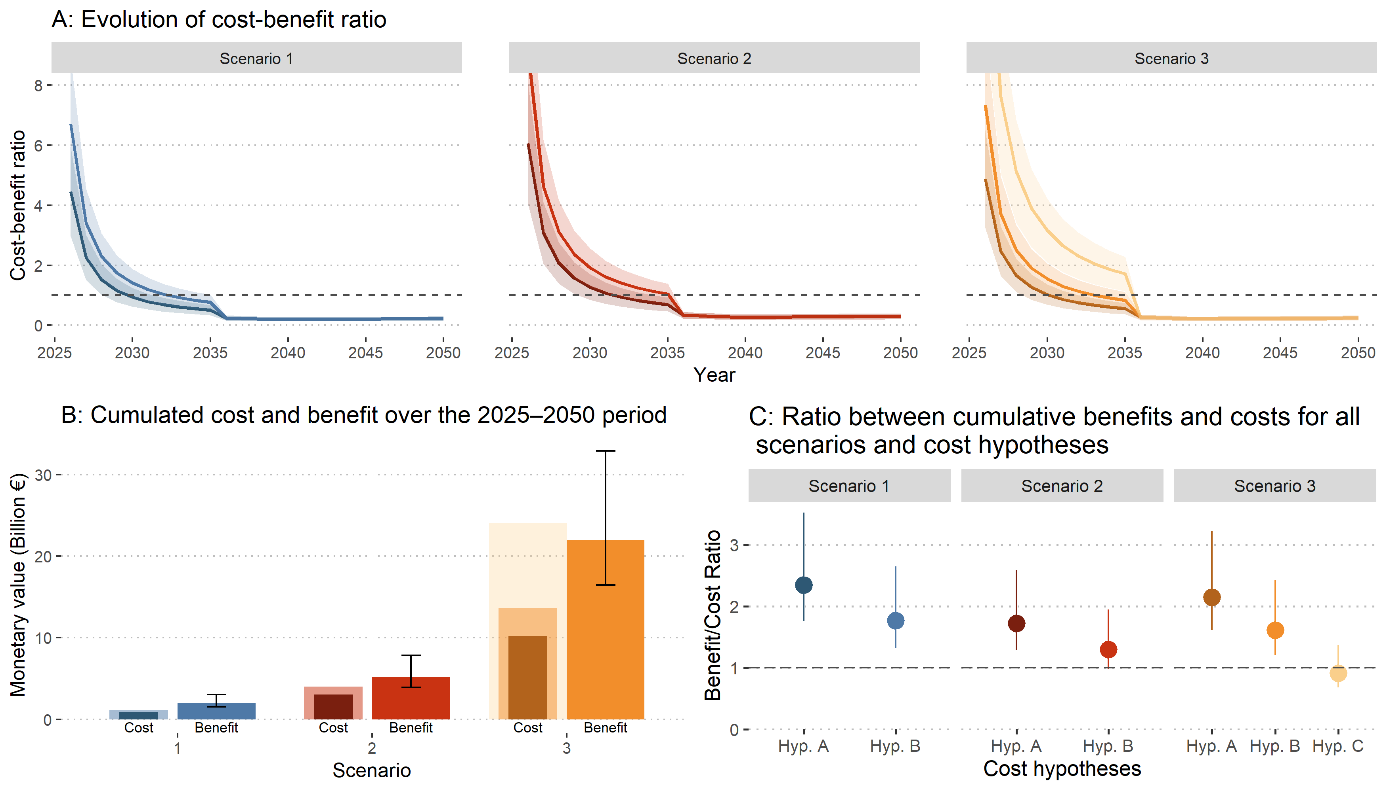

### Table S1: Technical path corresponding to hypotheses A, B and C.

| Steps | Techniques | **Hyp. A** | **Hyp. B** | **Hyp. C** |
| --- | --- | --- | --- | --- |
| 1. Preliminary analyses | Total - Preliminary analyses | **5** | **5** | **5** |
| 2. Building demolition | Asbestos removal | **0** | **0** | **1** |
|  | Demolition | **0** | **0** | **2** |
| 3. Desealing | Desealing by cutting | **0** | **0** | **0** |
|  | Desealing by scraping | **1,5** | **1,5** | **1,5** |
|  | Permeable paving installation | **1** | **1** | **1** |
|  | Gravel installation | **1** | **1** | **1** |
| 4. Soil remediation | Soil decontamination: excavation of polluted soils | **0** | **0** | **0** |
|  | Soil decontamination: excavation of polluted soils* | **0** | **0** | **0** |
|  | Soil decontamination: venting/bioventing | **0** | **0** | **0** |
|  | Soil decontamination: biopile | **0** | **0** | **0** |
|  | Soil decontamination: phytoextraction | **0** | **0** | **0** |
|  | Soil decontamination: myco-phytodegradation | **0** | **0** | **0** |
| 5. Soil improvement | In situ soil improvement | **1** | **0** | **0** |
|  | Soil construction | **0** | **1** | **1** |
|  | Earthworks | **0** | **1** | **1** |
|  | Soil decompaction | **1** | **0** | **0** |
|  | Incorporation of compost | **1** | **1** | **1** |
|  | Incorporation of topsoil | **0** | **1** | **1** |
|  | Bioaugmentation/biostimulation | **1** | **0** | **0** |
| 6. Management of waste and excavated soil | Disposal of non-hazardous waste | **0** | **0** | **0** |
|  | Storage of inert waste | **0** | **0** | **0** |
|  | Storage of hazardous waste | **0** | **0** | **0** |
| 7. Planting | Grass sowing | **0** | **0** | **0** |
|  | Planting of herbaceous vegetation | **2** | **2** | **2** |
|  | Planting of ligneous vegetation - sapling | **2** | **0** | **0** |
|  | Planting of ligneous vegetation - tree | **0** | **3** | **3** |
|  | Creation of water bodies | **0** | **0** | **0** |
| 8. Management and monitoring | Pollution monitoring | **0** | **0** | **0** |
|  | Ecological monitoring | **0** | **0** | **0** |
|  | Soil cycle maintenance | **2** | **0** | **0** |
|  | Maintenance of ligneous vegetation | **2** | **1,5** | **1,5** |
|  | Maintenance of herbaceous vegetation | **2** | **1,5** | **1,5** |

*The ‘0’ cells indicate a technique not used in the scenario. Certain cost items have been weighted upwards to reflect the specific constraints of dense urban environments such as Paris.*

### Table S2 : Event-study and average cumulative effect to estimate the calibration factor (output from ‘did_multiplegt_dyn’ package)

| Time | Estimate | Standard Error | Lower bound of the confidence interval | Upper bound of the confidence interval |
| --- | --- | --- | --- | --- |
| Effect_1 | 0,13725 | 0,06567 | 0,00855 | 0,26595 |
| Effect_2 | 0,09274 | 0,02174 | 0,05014 | 0,13535 |
| Effect_3 | 0,10219 | 0,01549 | 0,07182 | 0,13255 |
| Effect_4 | 0,12226 | 0,02439 | 0,07446 | 0,17006 |
| Effect_5 | 0,13968 | 0,01563 | 0,10906 | 0,17031 |
| Effect_6 | 0,08933 | 0,01607 | 0,05783 | 0,12082 |
| Effect_7 | 0,10984 | 0,01596 | 0,07856 | 0,14113 |
| Effect_8 | 0,09709 | 0,01589 | 0,06595 | 0,12823 |
| Effect_9 | 0,08777 | 0,01561 | 0,05718 | 0,11836 |
| Effect_10 | 0,0832 | 0,01572 | 0,05238 | 0,11401 |
| Effect_11 | 0,09499 | 0,0159 | 0,06382 | 0,12616 |
| Effect_12 | 0,10898 | 0,01808 | 0,07355 | 0,14442 |
| Effect_13 | 0,11267 | 0,01654 | 0,08026 | 0,14509 |
| Effect_14 | 0,12878 | 0,02716 | 0,07553 | 0,18202 |
| Total effect | 0,55513 | 0,0768 | 0,4046 | 0,70565 |

Average number of time periods over which a treatment effect is accumulated: 5.3741

CAUE, 2022. Les cours Oasis - Foire aux questions.

City of Paris, Direction de la voirie et des déplacements, 2024. Embellir votre quartier 12e : réunion publique, présentation du programme.

City of Paris, Direction de la voirie et des déplacements, 2023. Embellir votre quartier 13e : réunion publique, présentation du programme.

Cour des comptes, 2023. La politique de végétalisation de Paris, comme moyen d’adaptation au changement climatique : exercice 2017 et suivants.

De Chaisemartin, C., D’Haultfœuille, X., 2024. Difference-in-Differences Estimators of Intertemporal Treatment Effects. Review of Economics and Statistics 1–45. https://doi.org/10.1162/rest_a_01414

Salin, Charles Claron, Elodie Nguyen-Rabot, Nicolas Mondolfo, Harold Levrel, 2025. Les coûts de la restauration des sols urbains. CIRED Working Paper n°2024-96-FR. https://hal.science/hal-04904897

Salin, M., Claron, C., 2025. A dataset of cost estimates for urban soil restoration in France. https://doi.org/10.5281/ZENODO.15191255
